# Looking for crumbs in the obesity forest: anti-obesity interventions in the Mexican population. History, and systematic review with Meta-Analysis

**DOI:** 10.1101/2021.05.27.21257740

**Authors:** The Síntevi Group, Esperanza M. Garcia-Oropesa, Yoscelina E. Martinez-Lopez, Sonia María Ruiz-Cejudo, José Darío Martínez-Ezquerro, Alvaro Diaz-Badillo, Carlos Ramirez-Pfeiffer, Alejandra Bustamante-Fuentes, Elena B. Lopez-Sosa, Oscar O. Moctezuma-Chavez, Edna J. Nava-Gonzalez, Adriana L. Perales-Torres, Lucia M. Perez-Navarro, Marisol Rosas-Diaz, Kathleen Carter, Beatriz Tapia, Juan C. Lopez-Alvarenga

## Abstract

Mexicans and Mexican Americans share culture, genetic background, and predisposition for chronic complications associated with obesity and diabetes making imperative efficacious treatments and prevention. Obesity has been treated for centuries focused-on weight loss while other treatments on associated conditions like gout, diabetes (T2D), and hypertriglyceridemia. To date, there is no systematic review that synthetize the origin of obesity clinics in Mexico and the efforts to investigate treatments for obesity tested by randomized clinical trials (RCT).

We conducted systematic searches in Pubmed, Scopus, and Web of Science to retrieve anti-obesity RCT through 2019 and without inferior temporal limit. The systematic review included RCT of anti-obesity treatments in the Mexican adult population, including alternative medicine, pharmacological, nutritional, behavioral, and surgical interventions reporting biometric outcomes such as BMI, weight, waist circumference, triglycerides, glucose, among others. Studies with at least three months of treatment were included in the meta-analysis.

We found 634 entries, after removal of duplicates and screening the studies based on eligibility criteria, we analyzed 43, and 2 multinational-collaborative studies. Most of the national studies have small sample sizes, and the studied strategies do not have replications in the population. The nutrition/behavioral interventions were difficult to blind, and most studies have medium to high risk of bias. Nutritional/behavioral interventions and medications showed effects on BMI, waist circumference, and blood pressure. Simple measures like plain water instead of sweet beverages decrease triglycerides and systolic blood pressure. Participants with obesity and hypertension can have benefic effects with antioxidants, and treatment with insulin increase weight in those with T2D.

The study of obesity in Mexico has been on-going for more than four decades, but the interest on RCT just increased until this millennium, but with small sample sizes and lack of replication. The interventions affect different metabolic syndrome components, which should be analyzed in detail with the population living on the U.S.-Mexico border; therefore, bi-national collaboration is desirable to disentangle the cultural effects on this population’s treatment response.

## Introduction

Obesity phenotypes have been reproduced in arts and culture since the dawn of humanity. The Venus of Willendorf, made 25 to 30,000 years BCE during the upper Paleolithic and currently exhibited in the Natural History Museum in Vienna, is a figure depicting a woman with abdominal obesity symbolizing femininity, beauty, and fertility. In Mexico the artistic expression in the well-preserved Bonampak, which literally means painted wall, shows the Mayan ruling family of Bonampak, led by King Chaan Muwan and his wife Lady Rabbit (1). The scenes painted between 790 and 792 AD show human figures with signs of overweight or obesity. Other figurines from western Mexico show male abdominal distension, some of them may represent fat, ascites, cancer, or other underlying conditions with controversial significance (2).

For centuries, treatments for obesity have been described, mainly focused on weight loss and as a treatment for other secondary conditions such as gout and diabetes. In Greek and Roman art, obesity generated displeasure and sarcasm, being caricatured as an excess of alimentary and sexual stereotypes. This deviation from the norm was unacceptable in ancient Greek art, and they called by the term “Hippocratic Corpus” what we now call *morbid obesity*. The cause of obesity was considered the excess of fluids circulating in the body, therefore, the treatment consisted of restricting the fluid balance through diet, exercise, and medications. Since then, side effects of treatments have been described, like any leanness of the body causes corrugation of the skin, or if a woman is pregnant there would be risk of miscarriage.(3) The literature brought contrasting characters like Sancho Panza and Don Quixote, or Falstaff and Hamlet, stereotypes from Cervantes and Shakespeare, respectively. This jolly fat figure remains in traditional celebrations like the image of Santa Claus contrasting with slim Scrooge (4).

In the eighteenth-century, the medical literature documented the association of obesity with fatigue, gout, and breathing difficulties. Fat was considered reprehensible and medically undesirable. The progression of nutrition as science with direct calorimetry measurement experiments from Atwater and Benedict (5) between 1898 and 1900 analyzed the law of the conservation of energy applied to living organisms’ metabolism. They made a detailed description of demographic characteristics in men, diet, physical activity recorded in their “Metabolism experiments”. During the last century Vague (6) described the well-known android and gynecoid morphologies. He measured the perimeter and thickness of adipose tissue with calipers. Obesity has become a recognized clinical entity, subject of research and treatment with medication and behavioral techniques.

In the United States, Mexican Americans are considered part of the Hispanic Americans or Latino group. The U.S.-Mexico border represents this minority with active immigration, and a rapid increase in population. One of the Healthy People 2020 goals was to improve the health of all groups, requiring an understanding of the Hispanic culture, and health care needs for health promotion (7).

### Obesity in Mexico, a story never told

The Obesity Clinic started in 1959 at the Instituto Nacional de Nutricion Salvador Zubiran, by Dr. Luis Domenge, Dr. Carmen Ramos and Dr. Jorge Gonzalez-Barranco. Obesity was considered as an aesthetic but also a medical problem. The so-called epidemiologic transition, from infectious to chronic degenerative diseases, moved slowly from the 70s and 80s. The evolution of treating obesity as a medical problem was promoted by Dr. Gonzalez-Barranco based on scientific research and clinical trials with medications in the 90s.

The first attempt to classify obesity was using the Metropolitan Life Insurance Company (MLIC) which developed standard tables for “ideal” (MLIC 1942) and then “desirable” weight (MLIC 1959) based on the observed association of body weight with mortality. These standard tables were the platform for developing the current definition for underweight, normal, overweight, and obese individuals based on the body mass index (BMI) cut-offs (8).

The use of BMI as a reliable measurement started with the NHANES from 1988 to 2016. These studies demonstrated the age-adjusted prevalence of obesity in the United States increased progressively: from 22.9 to 39.6 percent. The main issue of concern in regard to BMI involves the growing obesity epidemic and the increasing population with high BMI numbers (9).

Since 1993 a series of population surveys were conducted systematically in Mexico using the BMI. The first National Survey on Chronic Diseases (ENEC from Spanish: Encuesta Nacional de Enfermedades Crónicas) highlighted obesity as a national public health problem. The prevalence of obesity in Mexico has increased substantially since the 1980s, and currently affects over 30% of the adult population (10). The epidemiological transition from undernourishment and infectious diseases to emergent chronic diseases were well documented in the ENEC. A sequel of undernourishment in presence of an obesogenic environment is homeorrhexis as an adaptive response to undernourishment. Homeorrexis or homeorhesis comes from the Greek homós, ’equal’; and rhéxis, ’violent rupture’, and refers to regulatory mechanisms that allow the body to change from one homeostatic, stable condition to another in a programmed fashion, e.g. growth during childhood or the onset of lactation (11). A combination of genetic and socioeconomic strata were conditions affecting stature. From North to South Mexico the ENEC data show a decrease in stature by expenses of the lower body segment (Figure 1), the sitting height is almost similar across regions. The stature can modify body composition despite BMI (12) and can be an indicator of socioeconomic inequality (13).

**Figure 1.**
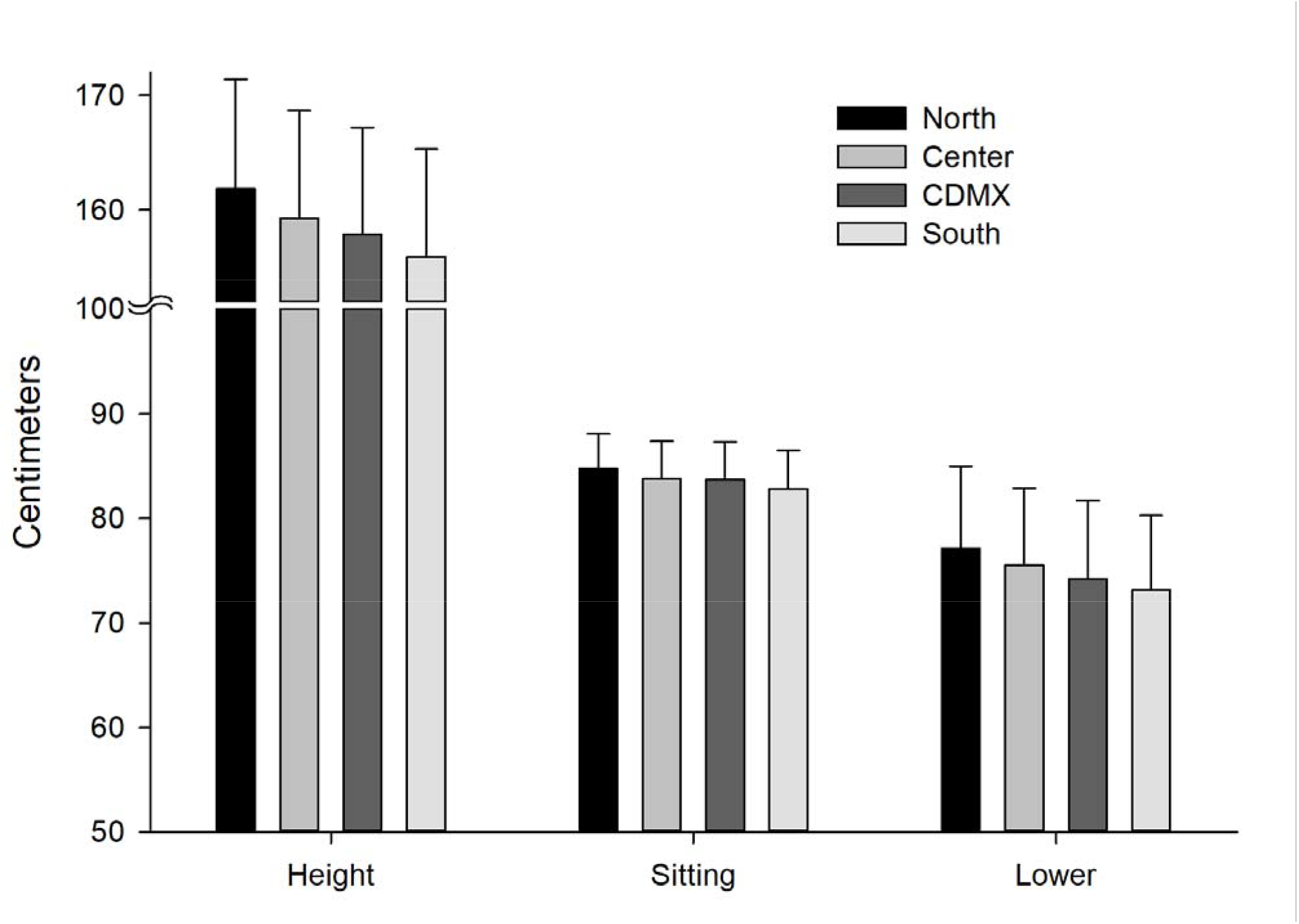
Height measurement standing up, sitting and the lower body segment. From North to South the height is lower at expenses to the lower segment of the body (p < 0.0001 adjusted by Bonferroni for all regions). The sitting height reflects the upper segment body and shows small differences between regions. Mean and standard deviations. Data obtained from the ENEC 1993. CDMX: Mexico City, Sitting: sitting height, Lower: Lower segment of the body (Height-sitting height).

The First Obesity meeting in Mexico with the NAASO and the Pan-American Endocrine Meetings were held in Cancun in 1997. These meetings were a landmark achievement for the study of obesity in Mexico with the first NOM (Mexican Official Norm) for obesity management, published in 1998 (14). Since this new millennium, there has been a spread of interest in obesity in other hospitals and Mexican states. Close collaboration with the Diabetes Division at the University of Texas San Antonio Health Science Center (UTSAHSC) and the South Texas Diabetes and Obesity Institute (STDOI) at the UTRGV has been done since then.

### Importance of gathering scientific literature in Mexico

Mexican Americans are spread all over the United States, the National Health and Nutrition Examination Surveys 1988-1994 showed children aged 4 to 17 years who born abroad had significantly lower prevalence of overweight / obesity compared to Mexican American children born in the U.S. (PR = 0.77, 95% CI: 0.61, 0.96). In contrast, during 2005-2014, there was no evidence of a difference in overweight / obesity at birth (PR = 0.95; 95% CI: 0.84, 1.07) and no differences with newer immigrants (<5 years living in the U.S.) compared with those born in the U.S. (15).

Regarding the diet quality, Yoshida et al., (16) reported age differences in diet quality influenced by acculturation (customary adoption of a new culture): older Mexican Americans had higher scores in Healthy Eating Index (HEI) indicating a better diet quality. For vegetables, fruits, and proteins, middle- aged adults had higher scores compared to young adults. Concerning HEI components, a 1-unit increase of acculturation was associated with 10% to 20% lower odds of attaining better scores for vegetables, fruits, dairy, sodium, and empty calories in almost all ages.

### Medication research and current anti-obesity guidelines

Some pharmacokinetics determinants of many drugs depend on the body size; for instance, obesity modifies the volume of distribution, and drug clearance, probably due to increased activity of cytochrome P450 2E1 and possible modifications on tubular reabsorption (17).

However, not only biology can explain the variability on losing weight, other factors are associated with the feasibility of following medical recommendations affected by cultural environment. The importance of lifestyle was defined in early times of weight loss intervention but was debated by the use of medication.

Numerous international published guidelines for anti-obesity treatment consider the local disparities and cultural differences of each geographic region. The management of obesity relies on diverse medical specialists, health professionals and government decisions. Primary prevention of obesity is fundamental and requires policies for favoring spaces for physical activity and a healthy environment. Harmonization on treatment cannot be global but can help to tailor weight loss treatments, and metabolic improvement for prevention of complications.

Since 2000 guidelines from the former North American Association for Study of Obesity (nowadays The Obesity Society -TOS) and the NIH Working Group were mainly based on dietary therapy, physical activity, and behavior therapy, and guided on the appropriate use of pharmacological and surgical interventions. The weight loss recommendation was for patients with BMI >30 and those with BMI between 25 and less than 30 with two or more complications. They suggested that pharmacotherapy should be used only in the context of a treatment program with diet, physical activity, and behavior therapy. Once the guide was published, only two drugs were approved for weight loss, sibutramine and orlistat (18).

The European guidelines also made emphasis on lifestyle modifications including nutrition and physical activity. The goals are risk reduction (even with modest weight loss i.e. 5-10% of initial body weight), attention on waist circumference and management of complications. They increase the number of drug treatments for obesity approved by FDA (Food and Drug Administration) and EMA (European Medicines Agency): orlistat, lorcaserin (only for FDA), phentermine/topiramate (only for FDA), bupropion/naltrexone and liraglutide. They recommend drug discontinuation if the patient does not reach 5% loss of initial weight after 12 weeks of treatment. This guide discusses metabolic surgery focusing on metabolic effects as primary outcomes instead being limited to weight loss (19).

The Endocrine Society in 2015 published the guideline for pharmacological management of obesity (20) implementing diet, exercise, and behavioral modification and suggesting drugs may amplify adherence to behavior change, especially for patients with a clinical history of failure in non-medication treatments.

The nutritional health status in Mexico was affected by government policies, the first supermarket chains selling American processed food in Mexico started in the 1940s. The government eliminated the subsidy of corn tortillas in 1999 with the objective to improve competitiveness in the global economy. This action loaded in closedown local tortilla factories not able to compete. The transition epidemiology from infectious to chronic diseases was rampant in this period. In 2008 the import tariffs on maize, bean, sugar, and mill were eliminated. In response to the nutritional problems and increase in obesity, in 2010 the Ministries of Public Education and of Health published the General Guidelines for Dispensing or Distribution of Foods and Beverages at School Food Establishments (SFEs). After a mass media campaign to reduce consumption of high caloric food, the Mexican congress, in 2014, excised a tax on high energy dense food (21).

This study aims to perform a systematic review with meta-analysis to synthesize and evaluate the evidence of anti-obesity treatments performed in Mexican adults with overweight and obesity. These treatments can include pharmaceutical, behavioral, surgical, nutritional, and alternative interventions designed as controlled clinical trials, to compare results within and between interventions, and finally, to discuss these findings with Mexican American studies.

## Methods

### Protocol registration and search strategy

The protocol was registered in PROSPERO on 11/17/2020 and assigned the registry number CRD42020221436. The search strategies included Pubmed, Scopus, and Web of Science databases to obtain published literature up to 2019 to include randomized controlled clinical trials for obesity conducted in Mexico. To identify additional studies and gray literature, we contacted Medical Societies such as the Endocrinology Society from Mexico and researchers from academic institutions such as UNAM. For inclusion in the meta-analysis, all interventions had to be conducted for at least three months and report both baseline and final BMI. The query was focused on all interventions with overweight or obese participants who underwent weight loss treatment. We included nutritional/behavioral treatments, with knowledge that many of these interventions cannot be blinded, therefore we assessed the possibility of bias using the Grading of Recommendations Assessment, Development and Evaluation (GRADE) approach (22). Medications, alternative medicine and surgical interventions were included in the review finished in December 2020.

An example of a search strategy performed in Pubmed without time period limits:

((“obesity”[MeSH Terms] OR “obesity”[All Fields]) AND (“therapy”[Subheading] OR “therapy”[All Fields] OR “treatment”[All Fields] OR “therapeutics”[MeSH Terms] OR “therapeutics”[All Fields])) AND (“mexico”[MeSH Terms] OR “mexico”[All Fields]) AND Clinical Trial[ptyp]

### Eligibility criteria

The systematic review included Mexican adult overweight or obese participants in controlled clinical trials subjected to pharmaceutical, behavioral, surgical, nutritional, or alternative interventions. Weight loss was the primary or secondary outcomes. We included studies published in English or Spanish at any time, conducted in Mexican centers and multicentric international studies with Mexican participants. For inclusion in the meta-analysis, treatments had to be conducted for at least three months and indicate baseline and final BMI. When available, we analyzed metabolic syndrome components (i.e., serum concentration of glucose, HDL-C, triglycerides, systolic and diastolic blood pressure, and waist circumference). We followed these criteria for the articles’ peer-screening and conducted a third final group review to resolve disagreements. We contacted the corresponding authors to clarify doubts and obtain additional information when necessary.

### Studies selection

We recovered 634 studies from three databases: Pubmed (n=180), Scopus (n=238), and Web of Science (n=216). After eliminating duplicate studies and applying the eligibility criteria, 589 studies were eliminated. The flux of the analyzed studies is described in Figure 2. Of the 45 included studies data were extracted using the Cochrane tool and quality assessed with the Jadad scale. There were 45 studies included in the qualitative synthesis and 25 in the meta-analysis. Of the studies included in the qualitative synthesis, 55 interventions were described: 25 with medications, 27 with nutrition and exercise, and 3 with surgical treatment (Table 1).

**Figure 2.**
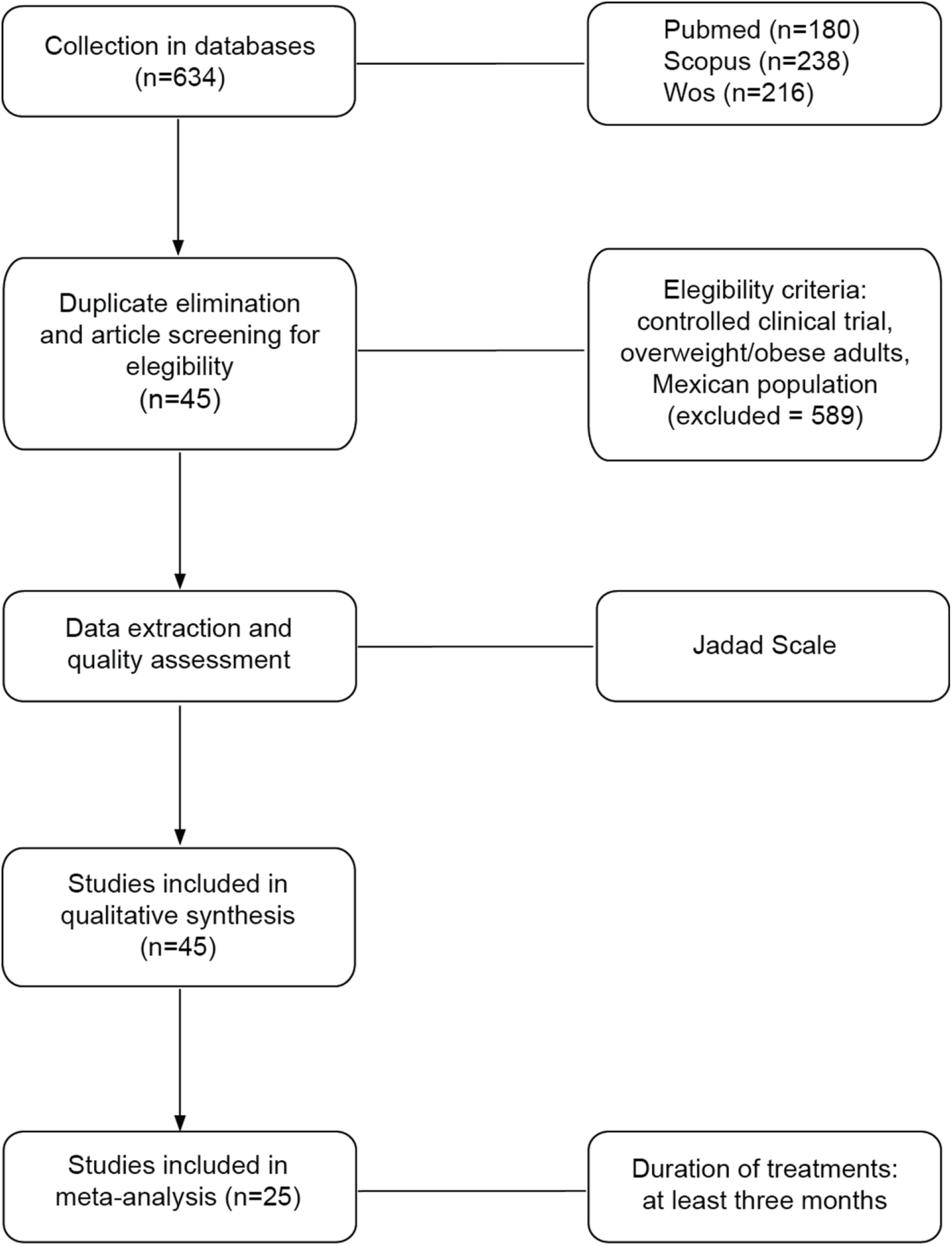
Procedure of the study. The flowchart shows the processes of collection, screening, quality assessment data extraction and analysis.

**Table 1.**
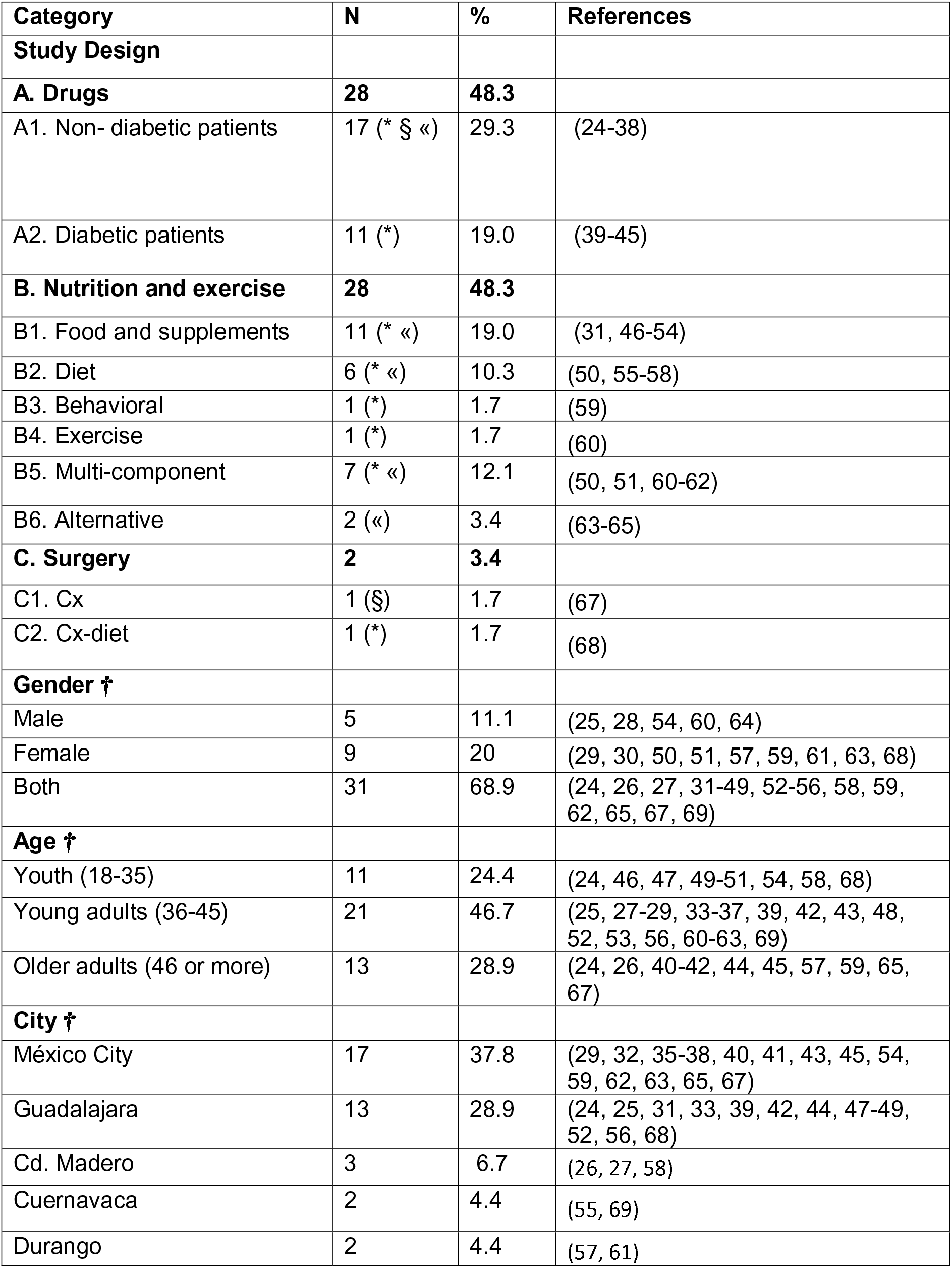
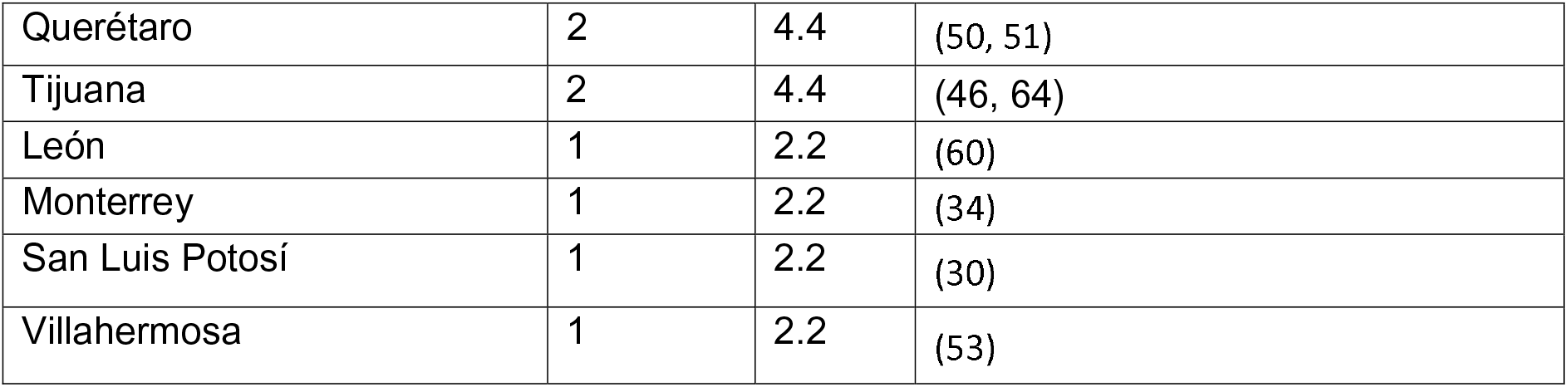
Characteristics of the analyzed interventions (n=58). Duration of study: («) < 3 months, (*) ≥ 3 months to 9 months, (§) ≥ 12 months. † Frequency from 45 analyzed studies.

| Category | N | % | References |
| --- | --- | --- | --- |
| **Study Design** |  |  |  |
| **A. Drugs** | **28** | **48.3** |  |
| A1. Non- diabetic patients | 17 (* § «) | 29.3 | (24-38) |
| A2. Diabetic patients | 11 (*) | 19.0 | (39-45) |
| **B. Nutrition and exercise** | **28** | **48.3** |  |
| B1. Food and supplements | 11 (* «) | 19.0 | (31, 46-54) |
| B2. Diet | 6 (* «) | 10.3 | (50, 55-58) |
| B3. Behavioral | 1 (*) | 1.7 | (59) |
| B4. Exercise | 1 (*) | 1.7 | (60) |
| B5. Multi-component | 7 (* «) | 12.1 | (50, 51, 60-62) |
| B6. Alternative | 2 («) | 3.4 | (63-65) |
| **C. Surgery** | **2** | **3.4** |  |
| C1. Cx | 1 (§) | 1.7 | (67) |
| C2. Cx-diet | 1 (*) | 1.7 | (68) |
| **Gender †** |  |  |  |
| Male | 5 | 11.1 | (25, 28, 54, 60, 64) |
| Female | 9 | 20 | (29, 30, 50, 51, 57, 59, 61, 63, 68) |
| Both | 31 | 68.9 | (24, 26, 27, 31-49, 52-56, 58, 59, 62, 65, 67, 69) |
| **Age †** |  |  |  |
| Youth (18-35) | 11 | 24.4 | (24, 46, 47, 49-51, 54, 58, 68) |
| Young adults (36-45) | 21 | 46.7 | (25, 27-29, 33-37, 39, 42, 43, 48, 52, 53, 56, 60-63, 69) |
| Older adults (46 or more) | 13 | 28.9 | (24, 26, 40-42, 44, 45, 57, 59, 65, 67) |
| **City †** |  |  |  |
| México City | 17 | 37.8 | (29, 32, 35-38, 40, 41, 43, 45, 54, 59, 62, 63, 65, 67) |
| Guadalajara | 13 | 28.9 | (24, 25, 31, 33, 39, 42, 44, 47-49, 52, 56, 68) |
| Cd. Madero | 3 | 6.7 | (26, 27, 58) |
| Cuernavaca | 2 | 4.4 | (55, 69) |
| Durango | 2 | 4.4 | (57, 61) |
| Querétaro | 2 | 4.4 | (50, 51) |
| Tijuana | 2 | 4.4 | (46, 64) |
| León | 1 | 2.2 | (60) |
| Monterrey | 1 | 2.2 | (34) |
| San Luis Potosí | 1 | 2.2 | (30) |
| Villahermosa | 1 | 2.2 | (53) |

### Data extraction process

The data extraction from the studies was done, by a team of 13 researchers, with a modified Cochrane tool for data collection form to obtain detailed information: type of intervention (drug, nutritional programs, behavioral treatments, use of drugs, surgical interventions or alternative medicine), age of intervention (childhood, adult), duration of treatments, year of the study development, sample size, groups of intervention, blindness of the treatment and the size of effects obtained in each study (Cohen’s d). Data extraction was performed in duplicate, and cases of discrepancy were re-analyzed in groups of 4 investigators. When it was necessary, authors were contacted to collect additional information. The main outcome was related with the reduction of BMI, waist circumference or percentage of body fat and biochemical parameters such as glucose, total cholesterol, triglycerides, HDL-c, blood pressure, HOMA-IR and Matsuda. WE used meta-regression to analyze the source of heterogeneity with mean age, mean BMI, location of the study (represented as latitude of the city of recruitment), sex distribution, and duration of the study. Adverse effects were also analyzed.

The quality assessment of the studies was done using the Jadad scale (23) and the risk of bias was assessed with GRADE checklist (22) with the following assessment guidelines:

Low risk studies were treatment with unpredictable allocation: A central office for allocation by phone, web, and pharmacy. Use of sequentially numbered, sealed, opaque envelopes. The drug containers are sequentially numbered and identical. Meanwhile high risk is predictable allocation, like staff know the random sequence in advance. Another high risk of bias was the use of envelopes or packaging without safeguards or non-random, predictable sequence. The attrition bias can be considered if there was a poor description on how much data was missing from each group, or the lack of reasons for missing data and how they were considered in the analysis. We were also interested in whether researchers used intention to treat analysis, imputation of missing values, or just per protocol analysis.

### Statistical analysis

The obtaining of sample size, means, and standard deviation was done with the data from the included studies. The summary of contrast was computed with Cohen’s-d differences. All models were analyzed with Restricted maximum likelihood (REML) random effects models, and the pooled effects were described with 95% confidence intervals. Heterogeneity was assessed with I^2 statistics, and we use meta-regression to analyze the heterogeneity. These statistical analyses were conducted with Stata 16.0 (StataCorp, College Station TX).

The network meta-analysis was computed for studies with medication only, because the designs of nutrition/behavior studies did not allow us to build nets. The analysis was performed with Stata 16.0 and CINeMA to define the network geometry, and effects comparisons. We did not have enough samples of studies to perform a rankogram.

## Results

We collected 634 studies from databases and after duplicate removal identified 64 controlled clinical trials from PubMed, 27 from Scopus and 15 from Web of Science conducted in Mexico.

### Studies characteristics

We included 45 anti-obesity national and multinational collaborative controlled clinical trials involving overweight and obese Mexican adults (>18 yr) subjected to distinct weight-loss interventions: pharmaceutical (25 studies), nutrition and behavioral (15 studies), surgical (2 studies), and alternative (3 studies) interventions (Table 1). Overall interventions included exclusively women were 15, men 5, and both sexes 35.

### Participant cities

Considering 45 interventions from 25 studies used in the quantitative analysis, Mexico City had the highest frequency of studies 38% (n=17), followed by Guadalajara 29% (n=13). The states close to the U.S. border were three (Nuevo Leon, Tamaulipas and Baja California). The details are described in Table 1.

### Risk of bias and quality

We performed a quality assessment at the intervention level. The Jadad mean value for nutritional/behaviour interventions was 3.6 (min 3, max 5), and for drug treatments was 3.7 (min 2, max 5). The nutrition/behaviour interventions had medium risk of bias (by GRADE) in 95% (n=18/19) and high risk of bias in 5% (n= 1/19). Physical activity was difficult to blind. The use of medication as intervention had very low risk of bias in 32% (n=7/22), medium 55% (n=12/22) and high risk in 14% (n=3/22). No differences in bias were found for intervention including T2D participants (Fisher’s exact test= 0.286).

### Synthesis of results

This meta-analysis included data from 2074 participants in nutrition/behaviour interventions and 5086 participants with medication, if we exclude multicentric international studies there were 1525 participants from studies exclusively made in Mexico. The main outcomes from individual studies are described in Tables 2 to 4. The forest plots with the pooled analysis are in Figures 3 to 10.

**Figure 3.**
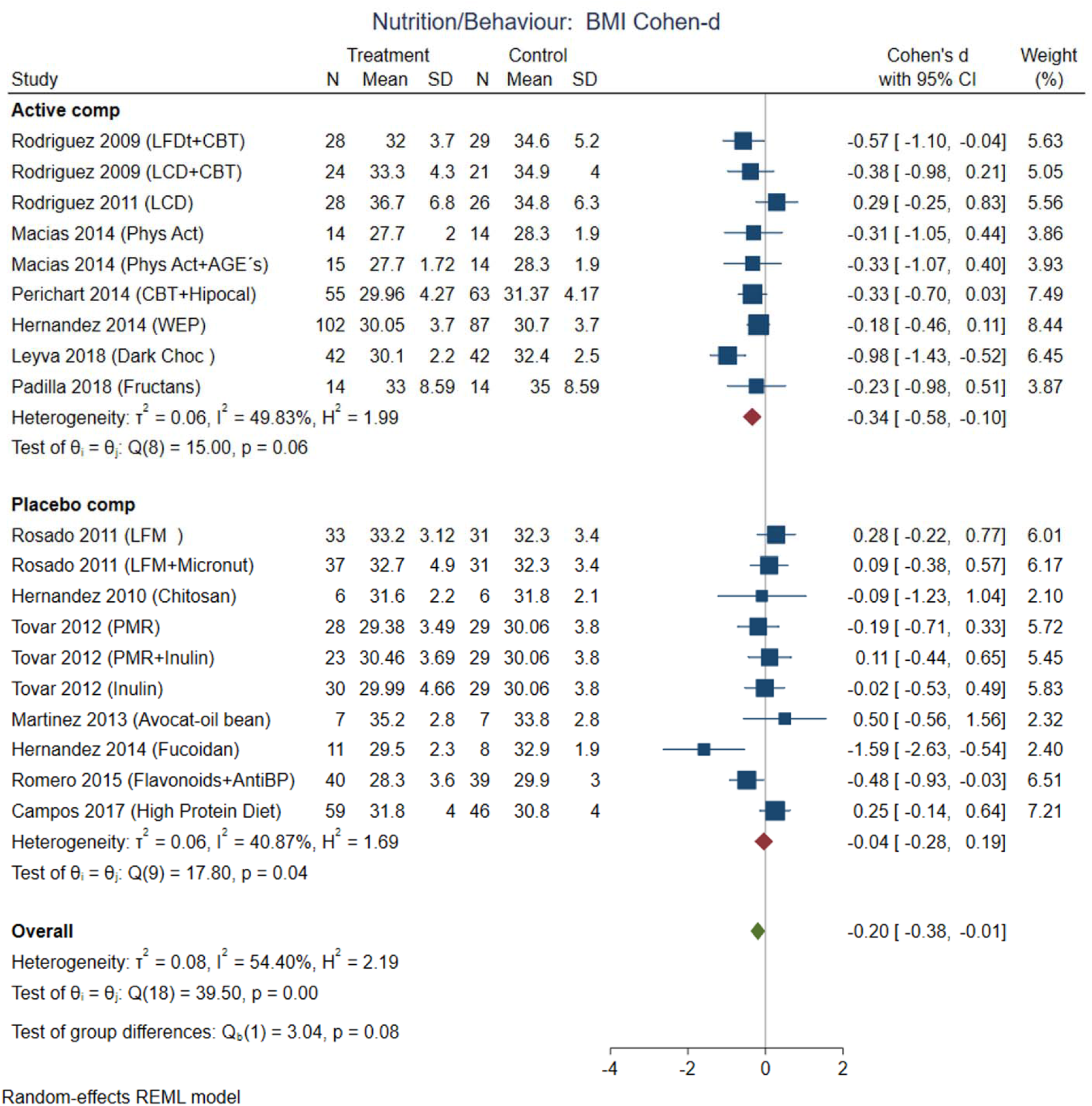
Pooled analysis of weighted size of effect by Cohen-d in BMI loss with nutritional and behavior interventions. The analysis was stratified by placebo or active comparator. LFDT: Low fat diet, LCD: Low carbohydrate diet, CBT: Cognitive-behavior therapy, Phys Act: Physical activity, Dark Choc: Dark chocolate, AGE: Advance glycation end-product, Hipocal: Hypocaloric diet, WEP: Water and Education Provision, LFM: Low fat milk, Micronut: Micronutrients, PMR: Partial meal replacement, AntiBP: Antihypertensive medication. REML: Restricted maximum likelihood.

**Table 2.**
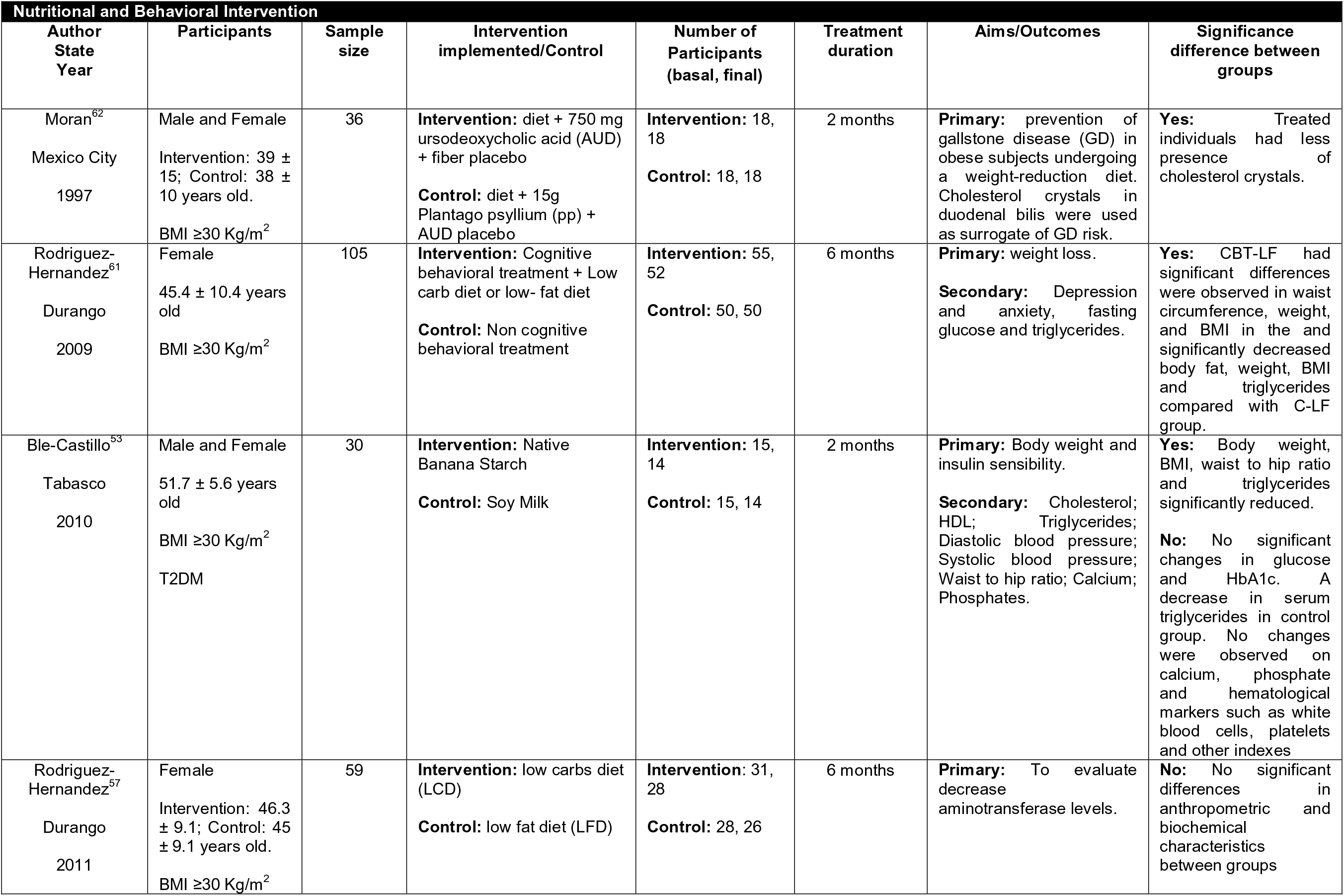
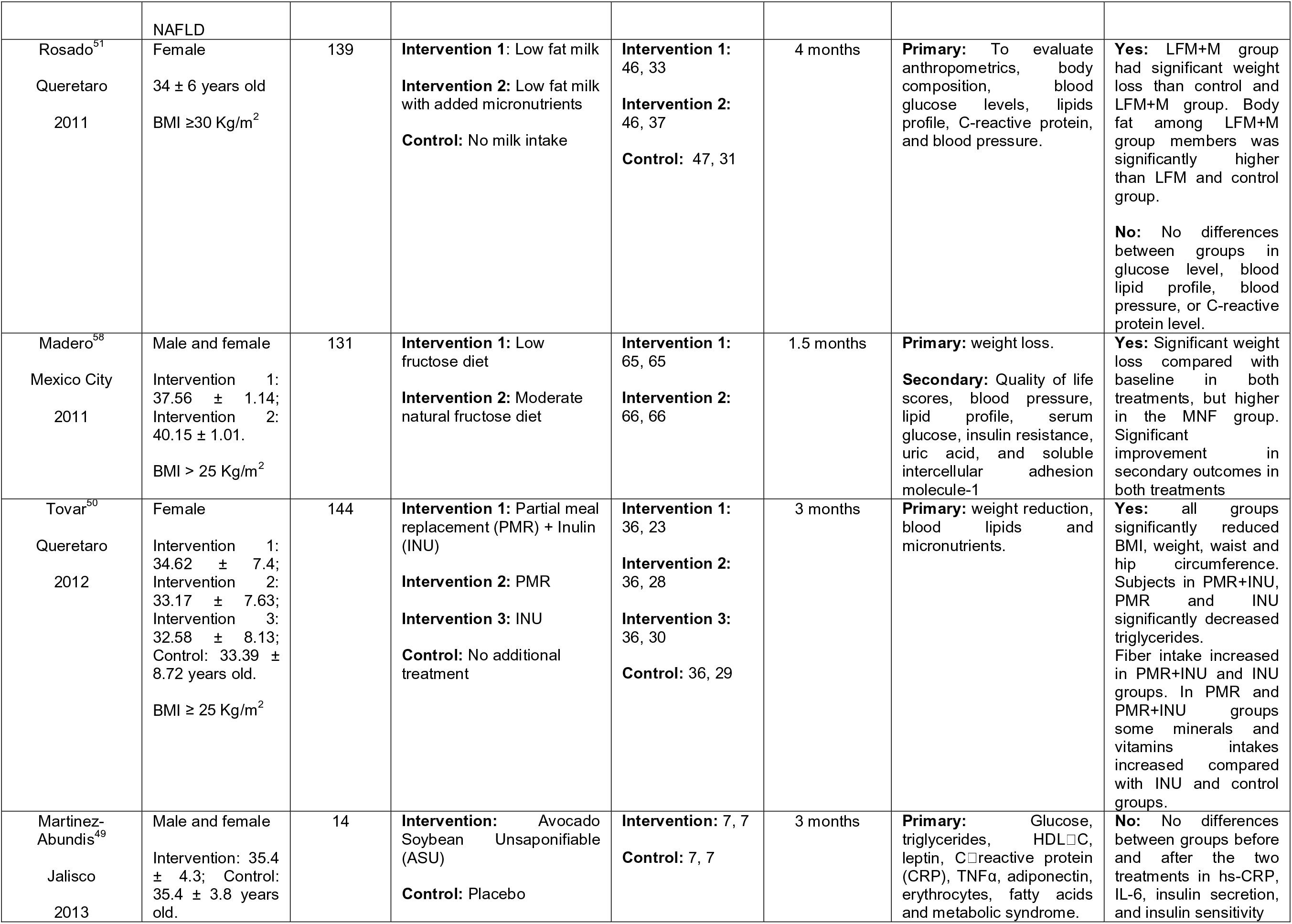
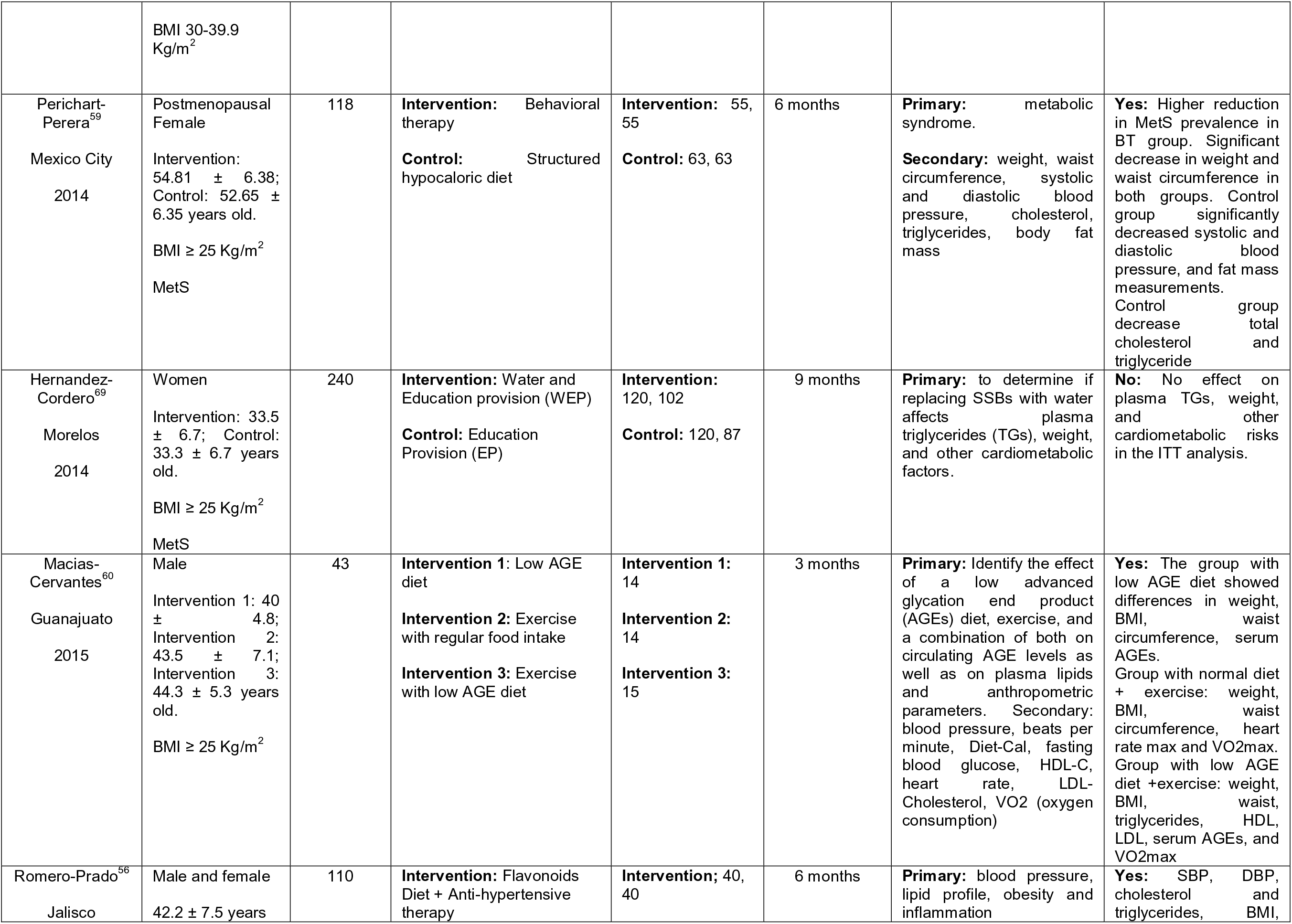
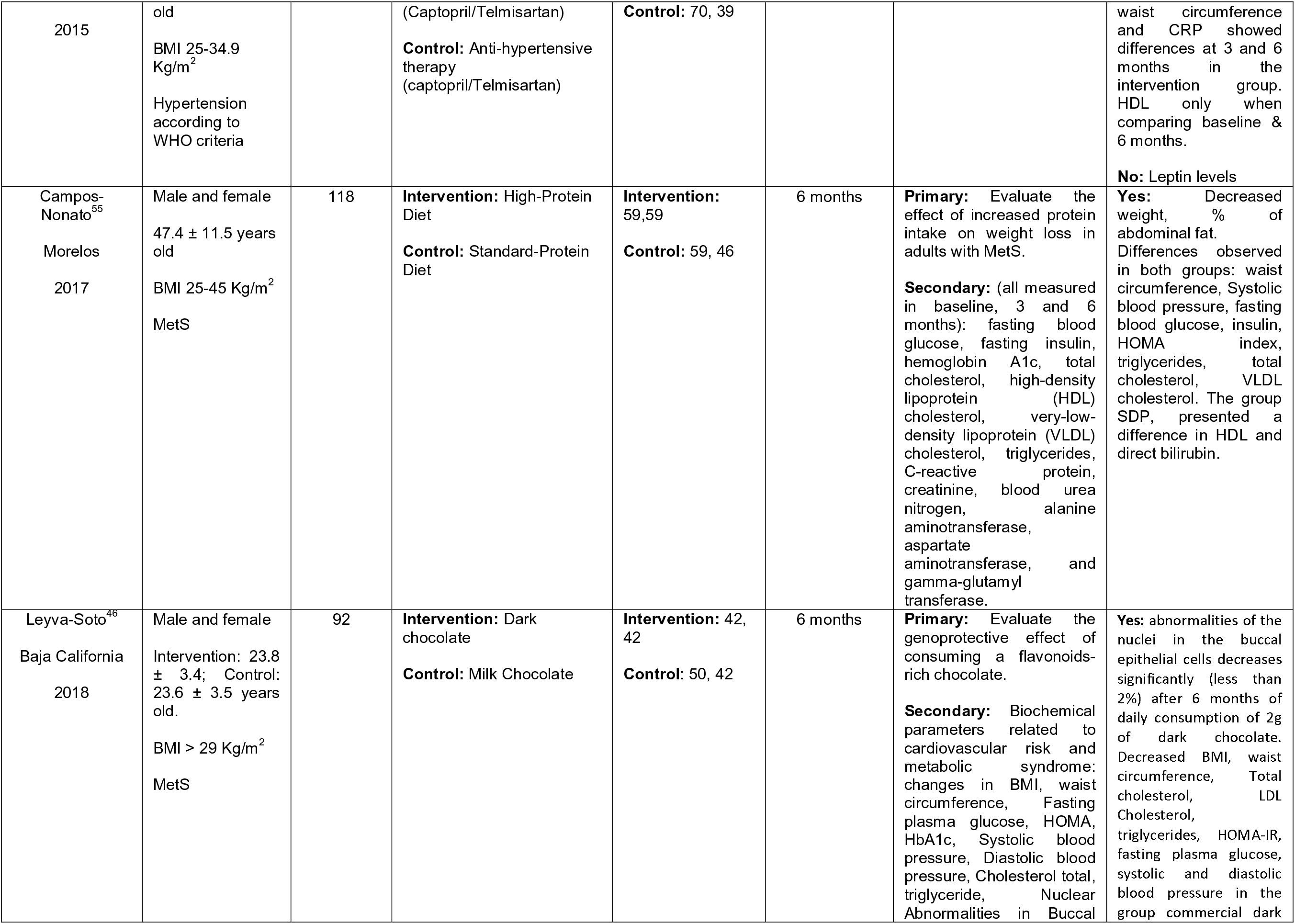
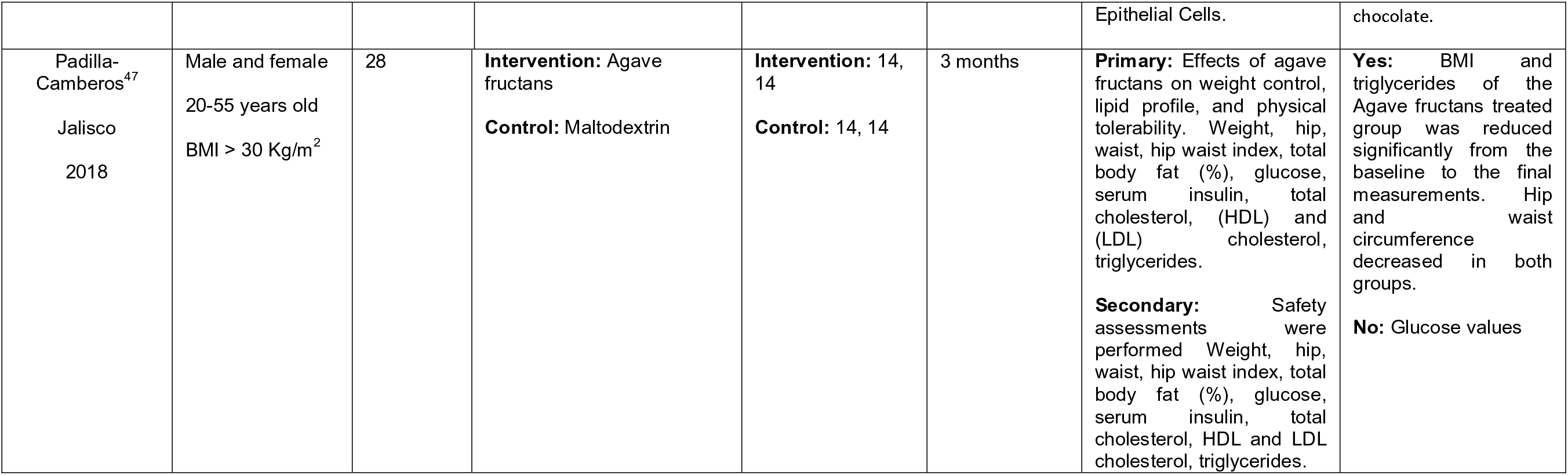
Descriptive characteristics and assessment of Nutrition/behavior interventions.

| Nutritional and Behavioral Intervention |  |  |  |  |  |  |  |
| --- | --- | --- | --- | --- | --- | --- | --- |
| Author<br>State<br>Year | Participants | Sample<br>size | Intervention<br>implemented/Control | Number of<br>Participants<br>(basal, final) | Treatment<br>duration | Aims/Outcomes | Significance<br>difference between<br>groups |
| Moran<sup>62</sup><br>Mexico City<br>1997 | Male and Female<br>Intervention: 39 ± 15; Control: 38 ± 10 years old.<br>BMI ≥30 Kg/m<sup>2</sup> | 36 | **Intervention:** diet + 750 mg ursodeoxycholic acid (AUD) + fiber placebo<br>**Control:** diet + 15g Plantago psyllium (pp) + AUD placebo | **Intervention:** 18, 18<br>**Control:** 18, 18 | 2 months | **Primary:** prevention of gallstone disease (GD) in obese subjects undergoing a weight-reduction diet. Cholesterol crystals in duodenal bilis were used as surrogate of GD risk. | **Yes:** Treated individuals had less presence of cholesterol crystals. |
| Rodriguez-Hernandez<sup>61</sup><br>Durango<br>2009 | Female<br>45.4 ± 10.4 years old<br>BMI ≥30 Kg/m<sup>2</sup> | 105 | **Intervention:** Cognitive behavioral treatment + Low carb diet or low- fat diet<br>**Control:** Non cognitive behavioral treatment | **Intervention:** 55, 52<br>**Control:** 50, 50 | 6 months | **Primary:** weight loss.<br>**Secondary:** Depression and anxiety, fasting glucose and triglycerides. | **Yes:** CBT-LF had significant differences were observed in waist circumference, weight, and BMI in the and significantly decreased body fat, weight, BMI and triglycerides compared with C-LF group. |
| Ble-Castillo<sup>53</sup><br>Tabasco<br>2010 | Male and Female<br>51.7 ± 5.6 years old<br>BMI ≥30 Kg/m<sup>2</sup><br>T2DM | 30 | **Intervention:** Native Banana Starch<br>**Control:** Soy Milk | **Intervention:** 15, 14<br>**Control:** 15, 14 | 2 months | **Primary:** Body weight and insulin sensibility.<br>**Secondary:** Cholesterol; HDL; Triglycerides; Diastolic blood pressure; Systolic blood pressure; Waist to hip ratio; Calcium; Phosphates. | **Yes:** Body weight, BMI, waist to hip ratio and triglycerides significantly reduced.<br>**No:** No significant changes in glucose and HbA1c. A decrease in serum triglycerides in control group. No changes were observed on calcium, phosphate and hematological markers such as white blood cells, platelets and other indexes |
| Rodriguez-Hernandez<sup>57</sup><br>Durango<br>2011 | Female<br>Intervention: 46.3 ± 9.1; Control: 45 ± 9.1 years old.<br>BMI ≥30 Kg/m<sup>2</sup> | 59 | **Intervention:** low carbs diet (LCD)<br>**Control:** low fat diet (LFD) | **Intervention:** 31, 28<br>**Control:** 28, 26 | 6 months | **Primary:** To evaluate decrease aminotransferase levels. | **No:** No significant differences in anthropometric and biochemical characteristics between groups |
|  | NAFLD |  |  |  |  |  |  |
| Rosado<sup>51</sup><br>Queretaro<br>2011 | Female<br>34 ± 6 years old<br>BMI ≥30 Kg/m<sup>2</sup> | 139 | **Intervention 1:** Low fat milk<br>**Intervention 2:** Low fat milk with added micronutrients<br>**Control:** No milk intake | **Intervention 1:** 46, 33<br>**Intervention 2:** 46, 37<br>**Control:** 47, 31 | 4 months | **Primary:** To evaluate anthropometrics, body composition, blood glucose levels, lipids profile, C-reactive protein, and blood pressure. | **Yes:** LFM+M group had significant weight loss than control and LFM+M group. Body fat among LFM+M group members was significantly higher than LFM and control group.<br>**No:** No differences between groups in glucose level, blood lipid profile, blood pressure, or C-reactive protein level. |
| Madero<sup>58</sup><br>Mexico City<br>2011 | Male and female<br>Intervention 1: 37.56 ± 1.14;<br>Intervention 2: 40.15 ± 1.01.<br>BMI > 25 Kg/m<sup>2</sup> | 131 | **Intervention 1:** Low fructose diet<br>**Intervention 2:** Moderate natural fructose diet | **Intervention 1:** 65, 65<br>**Intervention 2:** 66, 66 | 1.5 months | **Primary:** weight loss.<br>**Secondary:** Quality of life scores, blood pressure, lipid profile, serum glucose, insulin resistance, uric acid, and soluble intercellular adhesion molecule-1 | **Yes:** Significant weight loss compared with baseline in both treatments, but higher in the MNF group. Significant improvement in secondary outcomes in both treatments |
| Tovar<sup>50</sup><br>Queretaro<br>2012 | Female<br>Intervention 1: 34.62 ± 7.4;<br>Intervention 2: 33.17 ± 7.63;<br>Intervention 3: 32.58 ± 8.13;<br>Control: 33.39 ± 8.72 years old.<br>BMI ≥ 25 Kg/m<sup>2</sup> | 144 | **Intervention 1:** Partial meal replacement (PMR) + Inulin (INU)<br>**Intervention 2:** PMR<br>**Intervention 3:** INU<br>**Control:** No additional treatment | **Intervention 1:** 36, 23<br>**Intervention 2:** 36, 28<br>**Intervention 3:** 36, 30<br>**Control:** 36, 29 | 3 months | **Primary:** weight reduction, blood lipids and micronutrients. | **Yes:** all groups significantly reduced BMI, weight, waist and hip circumference. Subjects in PMR+INU, PMR and INU significantly decreased triglycerides. Fiber intake increased in PMR+INU and INU groups. In PMR and PMR+INU groups some minerals and vitamins intakes increased compared with INU and control groups. |
| Martinez-Abundis<sup>49</sup><br>Jalisco<br>2013 | Male and female<br>Intervention: 35.4 ± 4.3; Control: 35.4 ± 3.8 years old. | 14 | **Intervention:** Avocado Soybean Unsaponifiable (ASU)<br>**Control:** Placebo | **Intervention:** 7, 7<br>**Control:** 7, 7 | 3 months | **Primary:** Glucose, triglycerides, HDL-C, leptin, C-reactive protein (CRP), TNFα, adiponectin, erythrocytes, fatty acids and metabolic syndrome. | **No:** No differences between groups before and after the two treatments in hs-CRP, IL-6, insulin secretion, and insulin sensitivity |
|  | BMI 30-39.9<br>Kg/m<sup>2</sup> |  |  |  |  |  |  |
| Perichart-Perera<sup>59</sup><br>Mexico City<br>2014 | Postmenopausal Female<br>Intervention: 54.81 ± 6.38;<br>Control: 52.65 ± 6.35 years old.<br>BMI ≥ 25 Kg/m<sup>2</sup><br>MetS | 118 | **Intervention:** Behavioral therapy<br>**Control:** Structured hypocaloric diet | **Intervention:** 55, 55<br>**Control:** 63, 63 | 6 months | **Primary:** metabolic syndrome.<br>**Secondary:** weight, waist circumference, systolic and diastolic blood pressure, cholesterol, triglycerides, body fat mass | **Yes:** Higher reduction in MetS prevalence in BT group. Significant decrease in weight and waist circumference in both groups. Control group significantly decreased systolic and diastolic blood pressure, and fat mass measurements. Control group decrease total cholesterol and triglyceride |
| Hernandez-Cordero<sup>69</sup><br>Morelos<br>2014 | Women<br>Intervention: 33.5 ± 6.7; Control: 33.3 ± 6.7 years old.<br>BMI ≥ 25 Kg/m<sup>2</sup><br>MetS | 240 | **Intervention:** Water and Education provision (WEP)<br>**Control:** Education Provision (EP) | **Intervention:** 120, 102<br>**Control:** 120, 87 | 9 months | **Primary:** to determine if replacing SSBs with water affects plasma triglycerides (TGs), weight, and other cardiometabolic factors. | **No:** No effect on plasma TGs, weight, and other cardiometabolic risks in the ITT analysis. |
| Macias-Cervantes<sup>60</sup><br>Guanajuato<br>2015 | Male<br>Intervention 1: 40 ± 4.8;<br>Intervention 2: 43.5 ± 7.1;<br>Intervention 3: 44.3 ± 5.3 years old.<br>BMI ≥ 25 Kg/m<sup>2</sup> | 43 | **Intervention 1:** Low AGE diet<br>**Intervention 2:** Exercise with regular food intake<br>**Intervention 3:** Exercise with low AGE diet | **Intervention 1:** 14<br>**Intervention 2:** 14<br>**Intervention 3:** 15 | 3 months | **Primary:** Identify the effect of a low advanced glycation end product (AGEs) diet, exercise, and a combination of both on circulating AGE levels as well as on plasma lipids and anthropometric parameters. Secondary: blood pressure, beats per minute, Diet-Cal, fasting blood glucose, HDL-C, heart rate, LDL-Cholesterol, VO2 (oxygen consumption) | **Yes:** The group with low AGE diet showed differences in weight, BMI, waist circumference, serum AGEs. Group with normal diet + exercise: weight, BMI, waist circumference, heart rate max and VO2max. Group with low AGE diet + exercise: weight, BMI, waist, triglycerides, HDL, LDL, serum AGEs, and VO2max |
| Romero-Prado<sup>56</sup><br>Jalisco | Male and female<br>42.2 ± 7.5 years | 110 | **Intervention:** Flavonoids Diet + Anti-hypertensive therapy | **Intervention;** 40, 40 | 6 months | **Primary:** blood pressure, lipid profile, obesity and inflammation | **Yes:** SBP, DBP, cholesterol and triglycerides, BMI, |
| 2015 | old<br>BMI 25-34.9 Kg/m<sup>2</sup><br>Hypertension according to WHO criteria |  | (Captopril/Telmisartan)<br>**Control:** Anti-hypertensive therapy (captopril/Telmisartan) | **Control:** 70, 39 |  |  | waist circumference and CRP showed differences at 3 and 6 months in the intervention group. HDL only when comparing baseline & 6 months.<br>**No:** Leptin levels |
| Campos-Nonato<sup>55</sup><br>Morelos<br>2017 | Male and female<br>47.4 ± 11.5 years old<br>BMI 25-45 Kg/m<sup>2</sup><br>MetS | 118 | **Intervention:** High-Protein Diet<br>**Control:** Standard-Protein Diet | **Intervention:** 59,59<br>**Control:** 59, 46 | 6 months | **Primary:** Evaluate the effect of increased protein intake on weight loss in adults with MetS.<br>**Secondary:** (all measured in baseline, 3 and 6 months): fasting blood glucose, fasting insulin, hemoglobin A1c, total cholesterol, high-density lipoprotein (HDL) cholesterol, very-low-density lipoprotein (VLDL) cholesterol, triglycerides, C-reactive protein, creatinine, blood urea nitrogen, alanine aminotransferase, aspartate aminotransferase, and gamma-glutamyl transferase. | **Yes:** Decreased weight, % of abdominal fat. Differences observed in both groups: waist circumference, Systolic blood pressure, fasting blood glucose, insulin, HOMA index, triglycerides, total cholesterol, VLDL cholesterol. The group SDP, presented a difference in HDL and direct bilirubin. |
| Leyva-Soto<sup>46</sup><br>Baja California<br>2018 | Male and female<br>Intervention: 23.8 ± 3.4; Control: 23.6 ± 3.5 years old.<br>BMI > 29 Kg/m<sup>2</sup><br>MetS | 92 | **Intervention:** Dark chocolate<br>**Control:** Milk Chocolate | **Intervention:** 42, 42<br>**Control:** 50, 42 | 6 months | **Primary:** Evaluate the genoprotective effect of consuming a flavonoids-rich chocolate.<br>**Secondary:** Biochemical parameters related to cardiovascular risk and metabolic syndrome: changes in BMI, waist circumference, Fasting plasma glucose, HOMA, HbA1c, Systolic blood pressure, Diastolic blood pressure, Cholesterol total, triglyceride, Nuclear Abnormalities in Buccal | **Yes:** abnormalities of the nuclei in the buccal epithelial cells decreases significantly (less than 2%) after 6 months of daily consumption of 2g of dark chocolate. Decreased BMI, waist circumference, Total cholesterol, LDL Cholesterol, triglycerides, HOMA-IR, fasting plasma glucose, systolic and diastolic blood pressure in the group commercial dark |
|  |  |  |  |  |  | Epithelial Cells. | chocolate. |
| Padilla-Camberos<sup>47</sup><br>Jalisco<br>2018 | Male and female<br>20-55 years old<br>BMI > 30 Kg/m<sup>2</sup> | 28 | **Intervention:** Agave fructans<br>**Control:** Maltodextrin | **Intervention:** 14, 14<br>**Control:** 14, 14 | 3 months | **Primary:** Effects of agave fructans on weight control, lipid profile, and physical tolerability. Weight, hip, waist, hip waist index, total body fat (%), glucose, serum insulin, total cholesterol, (HDL) and (LDL) cholesterol, triglycerides.<br>**Secondary:** Safety assessments were performed Weight, hip, waist, hip waist index, total body fat (%), glucose, serum insulin, total cholesterol, HDL and LDL cholesterol, triglycerides. | **Yes:** BMI and triglycerides of the Agave fructans treated group was reduced significantly from the baseline to the final measurements. Hip and waist circumference decreased in both groups.<br>**No:** Glucose values |

**Table 3.**
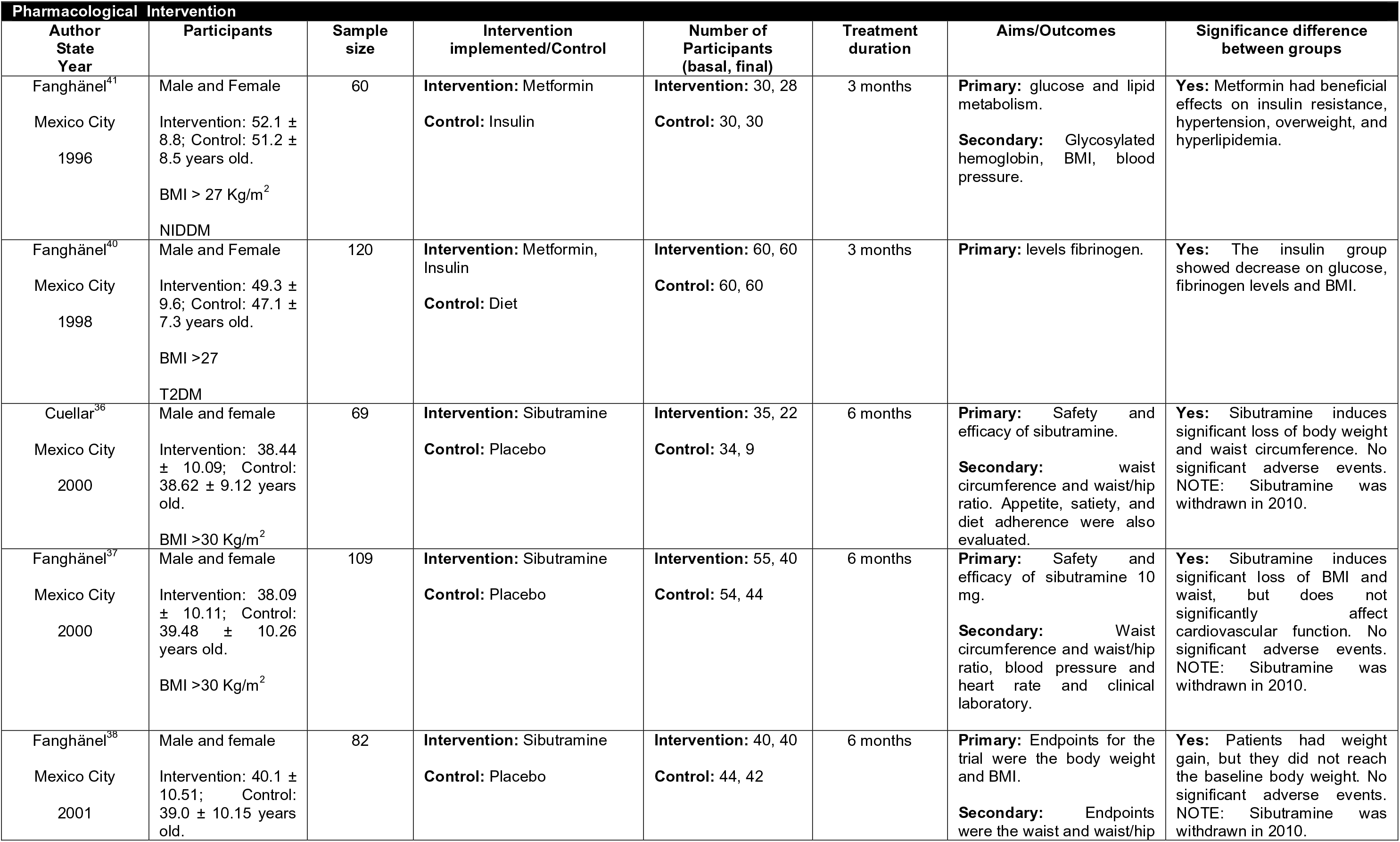
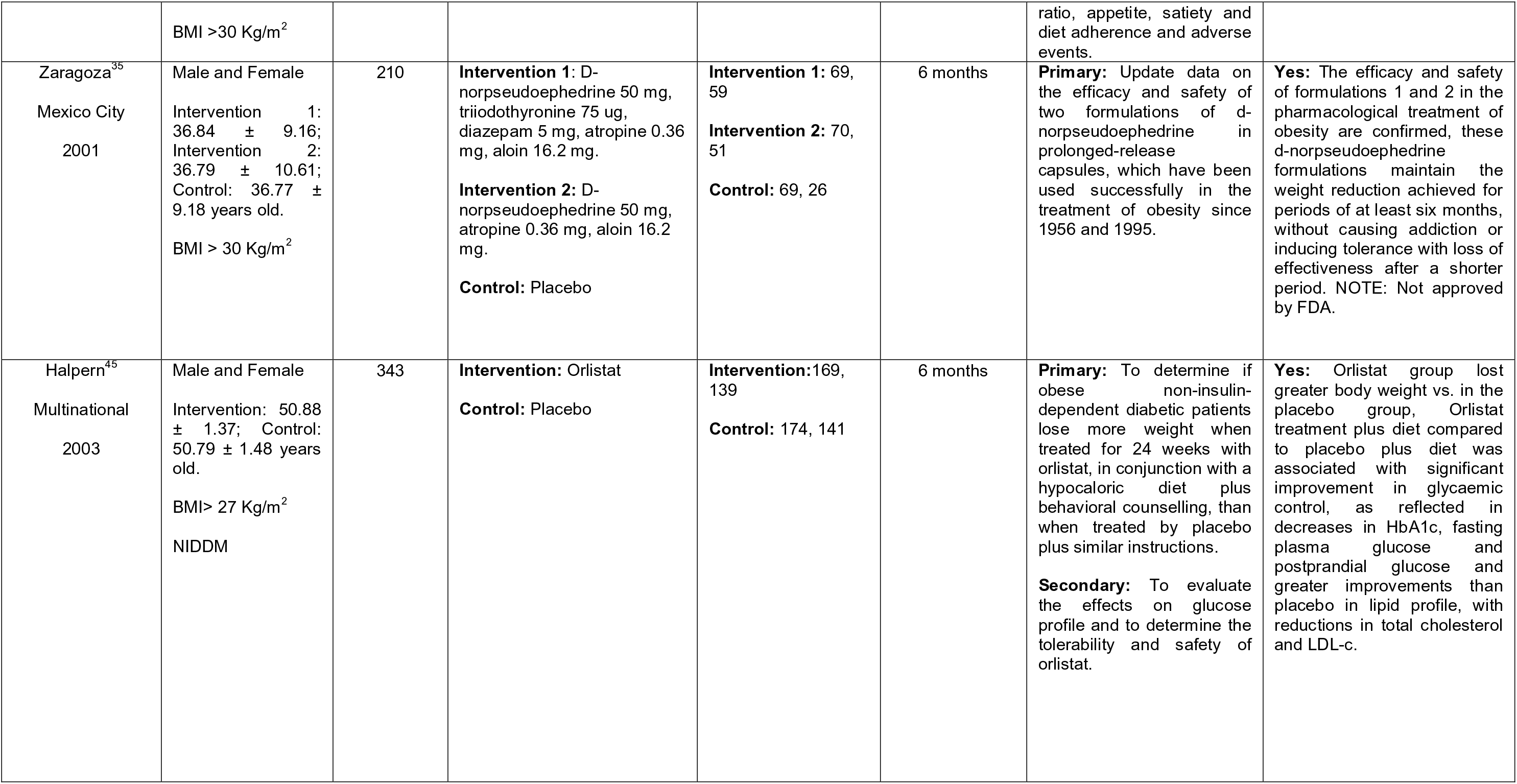
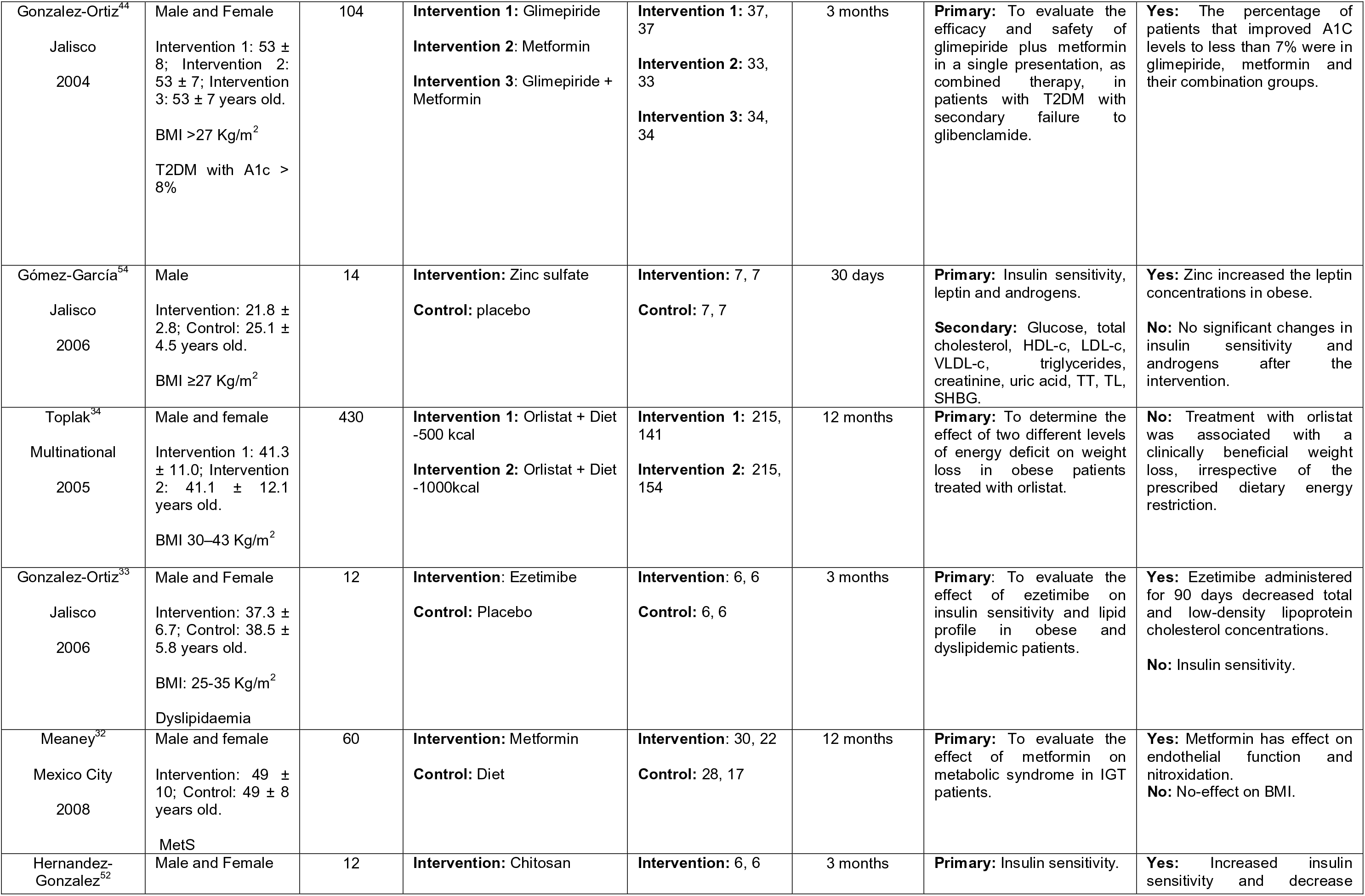
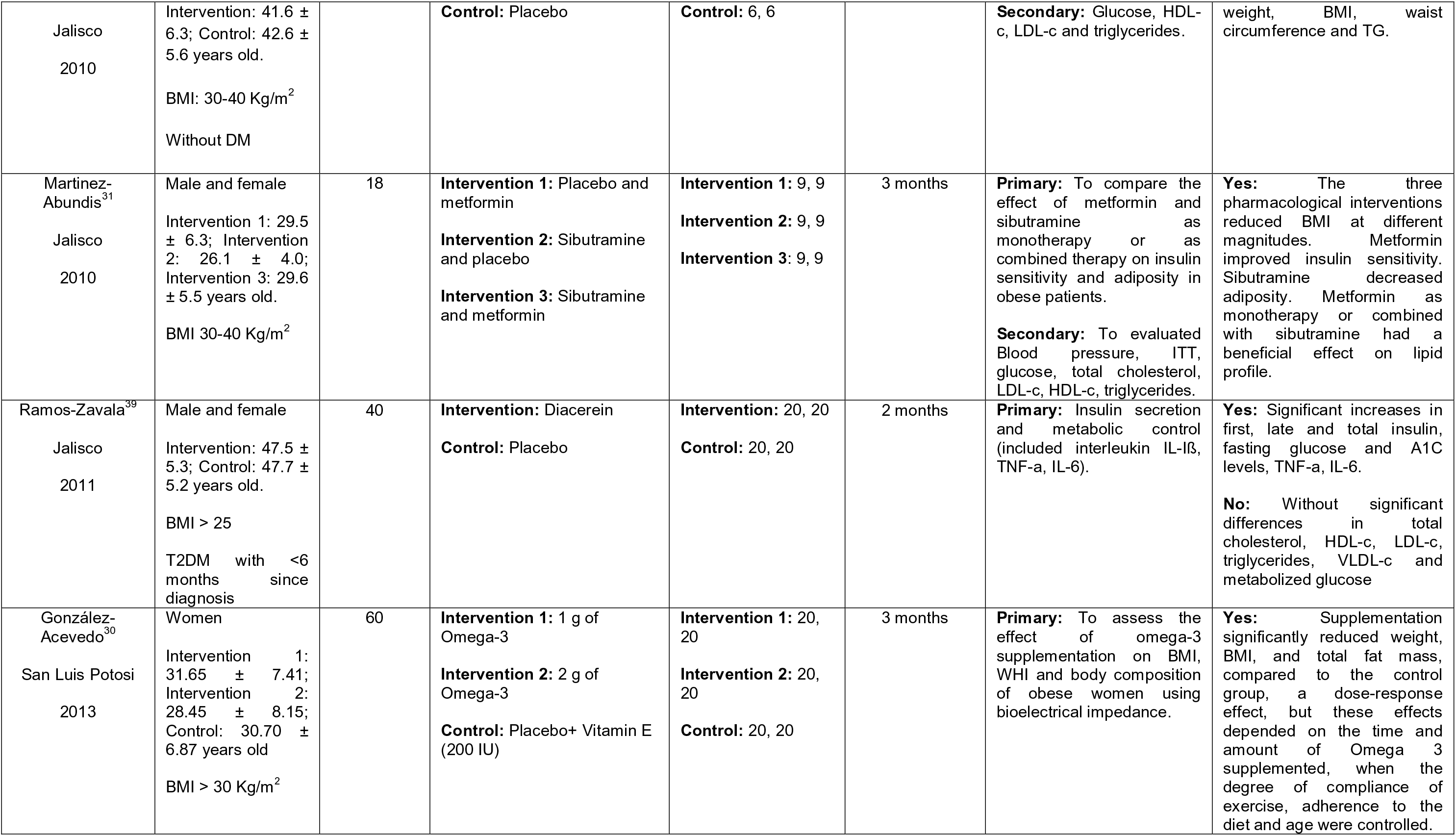
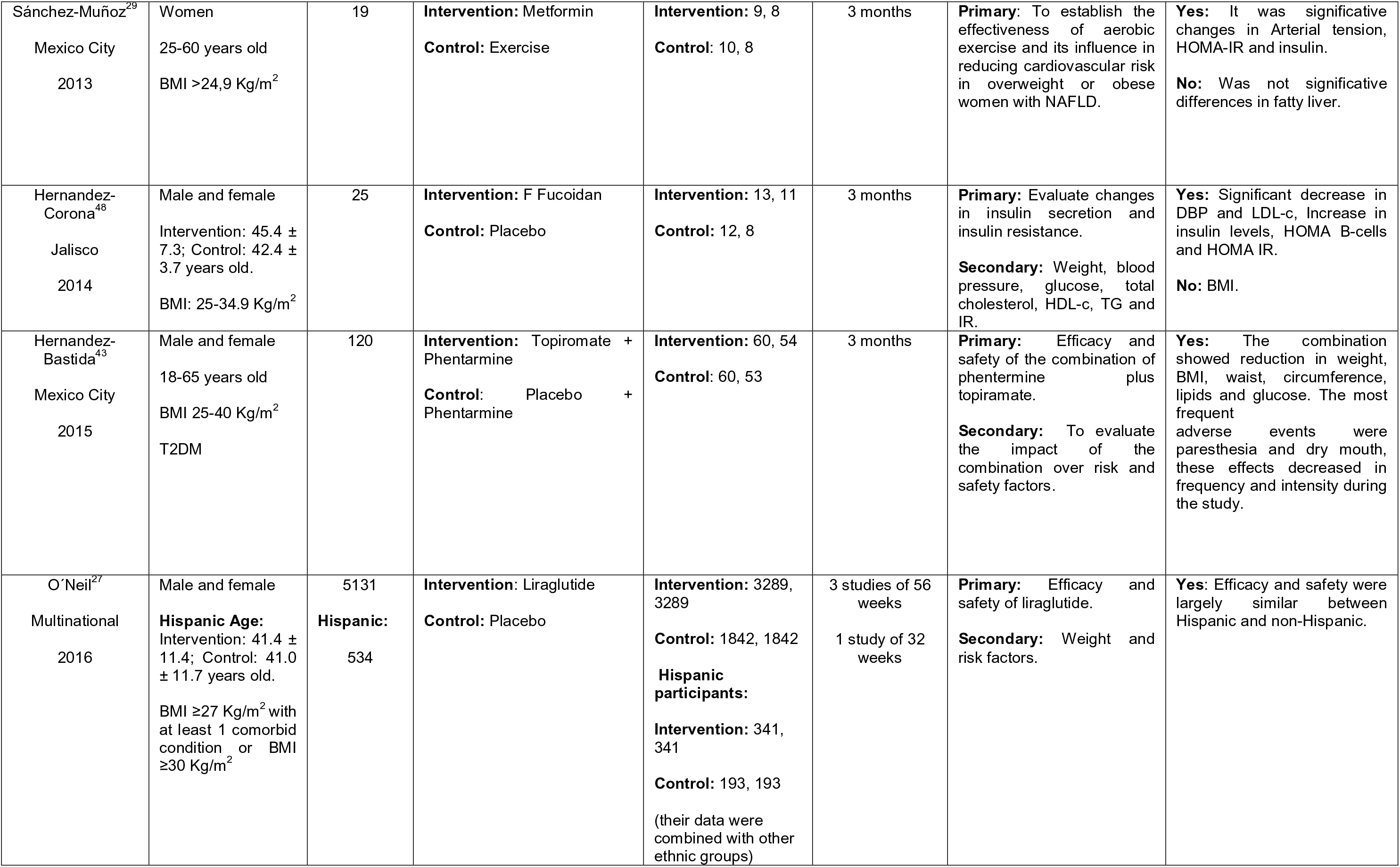
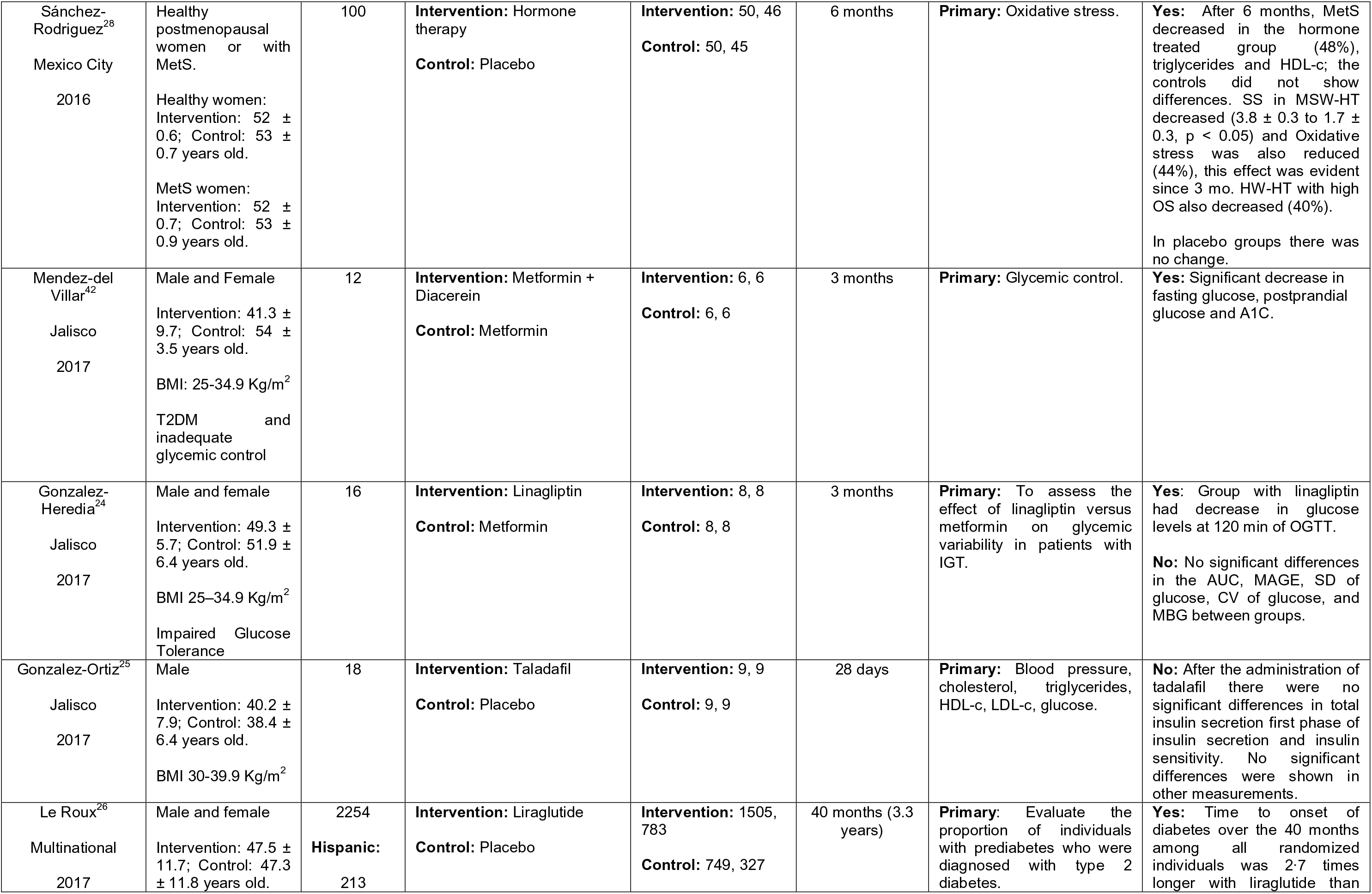
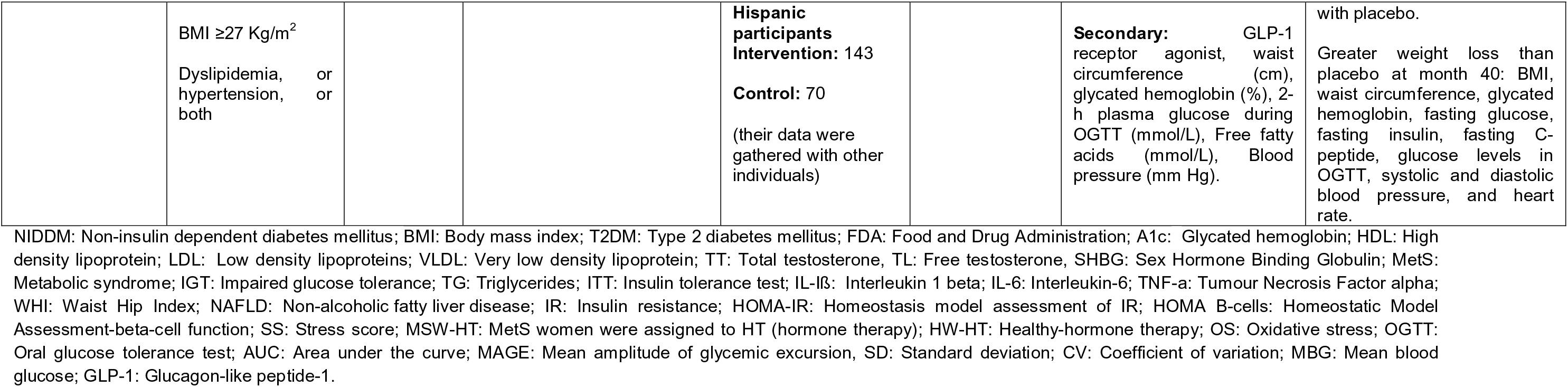
Descriptive characteristics and assessment of Medications.

| Pharmacological Intervention |  |  |  |  |  |  |  |
| --- | --- | --- | --- | --- | --- | --- | --- |
| Author<br>State<br>Year | Participants | Sample<br>size | Intervention<br>implemented/Control | Number of<br>Participants<br>(basal, final) | Treatment<br>duration | Aims/Outcomes | Significance difference<br>between groups |
| Fanghänel<sup>41</sup><br>Mexico City<br>1996 | Male and Female<br>Intervention: 52.1 ± 8.8; Control: 51.2 ± 8.5 years old.<br>BMI > 27 Kg/m<sup>2</sup><br>NIDDM | 60 | **Intervention:** Metformin<br>**Control:** Insulin | **Intervention:** 30, 28<br>**Control:** 30, 30 | 3 months | **Primary:** glucose and lipid metabolism.<br>**Secondary:** Glycosylated hemoglobin, BMI, blood pressure. | **Yes:** Metformin had beneficial effects on insulin resistance, hypertension, overweight, and hyperlipidemia. |
| Fanghänel<sup>40</sup><br>Mexico City<br>1998 | Male and Female<br>Intervention: 49.3 ± 9.6; Control: 47.1 ± 7.3 years old.<br>BMI >27<br>T2DM | 120 | **Intervention:** Metformin, Insulin<br>**Control:** Diet | **Intervention:** 60, 60<br>**Control:** 60, 60 | 3 months | **Primary:** levels fibrinogen. | **Yes:** The insulin group showed decrease on glucose, fibrinogen levels and BMI. |
| Cuellar<sup>36</sup><br>Mexico City<br>2000 | Male and female<br>Intervention: 38.44 ± 10.09; Control: 38.62 ± 9.12 years old.<br>BMI >30 Kg/m<sup>2</sup> | 69 | **Intervention:** Sibutramine<br>**Control:** Placebo | **Intervention:** 35, 22<br>**Control:** 34, 9 | 6 months | **Primary:** Safety and efficacy of sibutramine.<br>**Secondary:** waist circumference and waist/hip ratio. Appetite, satiety, and diet adherence were also evaluated. | **Yes:** Sibutramine induces significant loss of body weight and waist circumference. No significant adverse events. NOTE: Sibutramine was withdrawn in 2010. |
| Fanghänel<sup>37</sup><br>Mexico City<br>2000 | Male and female<br>Intervention: 38.09 ± 10.11; Control: 39.48 ± 10.26 years old.<br>BMI >30 Kg/m<sup>2</sup> | 109 | **Intervention:** Sibutramine<br>**Control:** Placebo | **Intervention:** 55, 40<br>**Control:** 54, 44 | 6 months | **Primary:** Safety and efficacy of sibutramine 10 mg.<br>**Secondary:** Waist circumference and waist/hip ratio, blood pressure and heart rate and clinical laboratory. | **Yes:** Sibutramine induces significant loss of BMI and waist, but does not significantly affect cardiovascular function. No significant adverse events. NOTE: Sibutramine was withdrawn in 2010. |
| Fanghänel<sup>38</sup><br>Mexico City<br>2001 | Male and female<br>Intervention: 40.1 ± 10.51; Control: 39.0 ± 10.15 years old. | 82 | **Intervention:** Sibutramine<br>**Control:** Placebo | **Intervention:** 40, 40<br>**Control:** 44, 42 | 6 months | **Primary:** Endpoints for the trial were the body weight and BMI.<br>**Secondary:** Endpoints were the waist and waist/hip | **Yes:** Patients had weight gain, but they did not reach the baseline body weight. No significant adverse events. NOTE: Sibutramine was withdrawn in 2010. |
|  | BMI >30 Kg/m<sup>2</sup> |  |  |  |  | ratio, appetite, satiety and diet adherence and adverse events. |  |
| Zaragoza<sup>35</sup><br>Mexico City<br>2001 | Male and Female<br>Intervention 1: 36.84 ± 9.16;<br>Intervention 2: 36.79 ± 10.61;<br>Control: 36.77 ± 9.18 years old.<br>BMI > 30 Kg/m<sup>2</sup> | 210 | **Intervention 1:** D-norpseudoephedrine 50 mg, triiodothyronine 75 ug, diazepam 5 mg, atropine 0.36 mg, aloin 16.2 mg.<br>**Intervention 2:** D-norpseudoephedrine 50 mg, atropine 0.36 mg, aloin 16.2 mg.<br>**Control:** Placebo | **Intervention 1:** 69, 59<br>**Intervention 2:** 70, 51<br>**Control:** 69, 26 | 6 months | **Primary:** Update data on the efficacy and safety of two formulations of d-norpseudoephedrine in prolonged-release capsules, which have been used successfully in the treatment of obesity since 1956 and 1995. | **Yes:** The efficacy and safety of formulations 1 and 2 in the pharmacological treatment of obesity are confirmed, these d-norpseudoephedrine formulations maintain the weight reduction achieved for periods of at least six months, without causing addiction or inducing tolerance with loss of effectiveness after a shorter period. NOTE: Not approved by FDA. |
| Halpern<sup>45</sup><br>Multinational<br>2003 | Male and Female<br>Intervention: 50.88 ± 1.37; Control: 50.79 ± 1.48 years old.<br>BMI > 27 Kg/m<sup>2</sup><br>NIDDM | 343 | **Intervention:** Orlistat<br>**Control:** Placebo | **Intervention:** 169, 139<br>**Control:** 174, 141 | 6 months | **Primary:** To determine if obese non-insulin-dependent diabetic patients lose more weight when treated for 24 weeks with orlistat, in conjunction with a hypocaloric diet plus behavioral counselling, than when treated by placebo plus similar instructions.<br>**Secondary:** To evaluate the effects on glucose profile and to determine the tolerability and safety of orlistat. | **Yes:** Orlistat group lost greater body weight vs. in the placebo group, Orlistat treatment plus diet compared to placebo plus diet was associated with significant improvement in glycaemic control, as reflected in decreases in HbA1c, fasting plasma glucose and postprandial glucose and greater improvements than placebo in lipid profile, with reductions in total cholesterol and LDL-c. |
| Gonzalez-Ortiz<sup>44</sup><br>Jalisco<br>2004 | Male and Female<br>Intervention 1: 53 ± 8; Intervention 2: 53 ± 7; Intervention 3: 53 ± 7 years old.<br>BMI >27 Kg/m<sup>2</sup><br>T2DM with A1c > 8% | 104 | **Intervention 1:** Glimepiride<br>**Intervention 2:** Metformin<br>**Intervention 3:** Glimepiride + Metformin | **Intervention 1:** 37, 37<br>**Intervention 2:** 33, 33<br>**Intervention 3:** 34, 34 | 3 months | **Primary:** To evaluate the efficacy and safety of glimepiride plus metformin in a single presentation, as combined therapy, in patients with T2DM with secondary failure to glibenclamide. | **Yes:** The percentage of patients that improved A1C levels to less than 7% were in glimepiride, metformin and their combination groups. |
| Gómez-García<sup>54</sup><br>Jalisco<br>2006 | Male<br>Intervention: 21.8 ± 2.8; Control: 25.1 ± 4.5 years old.<br>BMI ≥27 Kg/m<sup>2</sup> | 14 | **Intervention:** Zinc sulfate<br>**Control:** placebo | **Intervention:** 7, 7<br>**Control:** 7, 7 | 30 days | **Primary:** Insulin sensitivity, leptin and androgens.<br>**Secondary:** Glucose, total cholesterol, HDL-c, LDL-c, VLDL-c, triglycerides, creatinine, uric acid, TT, TL, SHBG. | **Yes:** Zinc increased the leptin concentrations in obese.<br>**No:** No significant changes in insulin sensitivity and androgens after the intervention. |
| Toplak<sup>34</sup><br>Multinational<br>2005 | Male and female<br>Intervention 1: 41.3 ± 11.0; Intervention 2: 41.1 ± 12.1 years old.<br>BMI 30–43 Kg/m<sup>2</sup> | 430 | **Intervention 1:** Orlistat + Diet -500 kcal<br>**Intervention 2:** Orlistat + Diet -1000kcal | **Intervention 1:** 215, 141<br>**Intervention 2:** 215, 154 | 12 months | **Primary:** To determine the effect of two different levels of energy deficit on weight loss in obese patients treated with orlistat. | **No:** Treatment with orlistat was associated with a clinically beneficial weight loss, irrespective of the prescribed dietary energy restriction. |
| Gonzalez-Ortiz<sup>33</sup><br>Jalisco<br>2006 | Male and Female<br>Intervention: 37.3 ± 6.7; Control: 38.5 ± 5.8 years old.<br>BMI: 25-35 Kg/m<sup>2</sup><br>Dyslipidaemia | 12 | **Intervention:** Ezetimibe<br>**Control:** Placebo | **Intervention:** 6, 6<br>**Control:** 6, 6 | 3 months | **Primary:** To evaluate the effect of ezetimibe on insulin sensitivity and lipid profile in obese and dyslipidemic patients. | **Yes:** Ezetimibe administered for 90 days decreased total and low-density lipoprotein cholesterol concentrations.<br>**No:** Insulin sensitivity. |
| Meaney<sup>32</sup><br>Mexico City<br>2008 | Male and female<br>Intervention: 49 ± 10; Control: 49 ± 8 years old.<br>MetS | 60 | **Intervention:** Metformin<br>**Control:** Diet | **Intervention:** 30, 22<br>**Control:** 28, 17 | 12 months | **Primary:** To evaluate the effect of metformin on metabolic syndrome in IGT patients. | **Yes:** Metformin has effect on endothelial function and nitrooxidation.<br>**No:** No-effect on BMI. |
| Hernandez-Gonzalez<sup>52</sup> | Male and Female | 12 | **Intervention:** Chitosan | **Intervention:** 6, 6 | 3 months | **Primary:** Insulin sensitivity. | **Yes:** Increased insulin sensitivity and decrease |
| Jalisco<br>2010 | Intervention: 41.6 ± 6.3; Control: 42.6 ± 5.6 years old.<br>BMI: 30-40 Kg/m<sup>2</sup><br>Without DM |  | **Control:** Placebo | **Control:** 6, 6 |  | **Secondary:** Glucose, HDL-c, LDL-c and triglycerides. | weight, BMI, waist circumference and TG. |
| Martinez-Abundis<sup>31</sup><br>Jalisco<br>2010 | Male and female<br>Intervention 1: 29.5 ± 6.3; Intervention 2: 26.1 ± 4.0; Intervention 3: 29.6 ± 5.5 years old.<br>BMI 30-40 Kg/m<sup>2</sup> | 18 | **Intervention 1:** Placebo and metformin<br>**Intervention 2:** Sibutramine and placebo<br>**Intervention 3:** Sibutramine and metformin | **Intervention 1:** 9, 9<br>**Intervention 2:** 9, 9<br>**Intervention 3:** 9, 9 | 3 months | **Primary:** To compare the effect of metformin and sibutramine as monotherapy or as combined therapy on insulin sensitivity and adiposity in obese patients.<br>**Secondary:** To evaluated Blood pressure, ITT, glucose, total cholesterol, LDL-c, HDL-c, triglycerides. | **Yes:** The three pharmacological interventions reduced BMI at different magnitudes. Metformin improved insulin sensitivity. Sibutramine decreased adiposity. Metformin as monotherapy or combined with sibutramine had a beneficial effect on lipid profile. |
| Ramos-Zavala<sup>39</sup><br>Jalisco<br>2011 | Male and female<br>Intervention: 47.5 ± 5.3; Control: 47.7 ± 5.2 years old.<br>BMI > 25<br>T2DM with <6 months since diagnosis | 40 | **Intervention:** Diacerein<br>**Control:** Placebo | **Intervention:** 20, 20<br>**Control:** 20, 20 | 2 months | **Primary:** Insulin secretion and metabolic control (included interleukin IL-1β, TNF-α, IL-6). | **Yes:** Significant increases in first, late and total insulin, fasting glucose and A1C levels, TNF-α, IL-6.<br>**No:** Without significant differences in total cholesterol, HDL-c, LDL-c, triglycerides, VLDL-c and metabolized glucose |
| González-Acevedo<sup>30</sup><br>San Luis Potosi<br>2013 | Women<br>Intervention 1: 31.65 ± 7.41; Intervention 2: 28.45 ± 8.15; Control: 30.70 ± 6.87 years old<br>BMI > 30 Kg/m<sup>2</sup> | 60 | **Intervention 1:** 1 g of Omega-3<br>**Intervention 2:** 2 g of Omega-3<br>**Control:** Placebo+ Vitamin E (200 IU) | **Intervention 1:** 20, 20<br>**Intervention 2:** 20, 20<br>**Control:** 20, 20 | 3 months | **Primary:** To assess the effect of omega-3 supplementation on BMI, WHI and body composition of obese women using bioelectrical impedance. | **Yes:** Supplementation significantly reduced weight, BMI, and total fat mass, compared to the control group, a dose-response effect, but these effects depended on the time and amount of Omega 3 supplemented, when the degree of compliance of exercise, adherence to the diet and age were controlled. |
| Sánchez-Muñoz<sup>29</sup><br>Mexico City<br>2013 | Women<br>25-60 years old<br>BMI >24,9 Kg/m<sup>2</sup> | 19 | **Intervention:** Metformin<br>**Control:** Exercise | **Intervention:** 9, 8<br>**Control:** 10, 8 | 3 months | **Primary:** To establish the effectiveness of aerobic exercise and its influence in reducing cardiovascular risk in overweight or obese women with NAFLD. | **Yes:** It was significant changes in Arterial tension, HOMA-IR and insulin.<br>**No:** Was not significant differences in fatty liver. |
| Hernandez-Corona<sup>48</sup><br>Jalisco<br>2014 | Male and female<br>Intervention: 45.4 ± 7.3; Control: 42.4 ± 3.7 years old.<br>BMI: 25-34.9 Kg/m<sup>2</sup> | 25 | **Intervention:** F Fucoidan<br>**Control:** Placebo | **Intervention:** 13, 11<br>**Control:** 12, 8 | 3 months | **Primary:** Evaluate changes in insulin secretion and insulin resistance.<br>**Secondary:** Weight, blood pressure, glucose, total cholesterol, HDL-c, TG and IR. | **Yes:** Significant decrease in DBP and LDL-c, Increase in insulin levels, HOMA B-cells and HOMA IR.<br>**No:** BMI. |
| Hernandez-Bastida<sup>43</sup><br>Mexico City<br>2015 | Male and female<br>18-65 years old<br>BMI 25-40 Kg/m<sup>2</sup><br>T2DM | 120 | **Intervention:** Topiramate + Phentarmine<br>**Control:** Placebo + Phentarmine | **Intervention:** 60, 54<br>**Control:** 60, 53 | 3 months | **Primary:** Efficacy and safety of the combination of phentermine plus topiramate.<br>**Secondary:** To evaluate the impact of the combination over risk and safety factors. | **Yes:** The combination showed reduction in weight, BMI, waist, circumference, lipids and glucose. The most frequent adverse events were paresthesia and dry mouth, these effects decreased in frequency and intensity during the study. |
| O'Neil<sup>27</sup><br>Multinational<br>2016 | Male and female<br>**Hispanic Age:**<br>Intervention: 41.4 ± 11.4; Control: 41.0 ± 11.7 years old.<br>BMI ≥27 Kg/m<sup>2</sup> with at least 1 comorbid condition or BMI ≥30 Kg/m<sup>2</sup> | 5131<br>**Hispanic:**<br>534 | **Intervention:** Liraglutide<br>**Control:** Placebo | **Intervention:** 3289, 3289<br>**Control:** 1842, 1842<br>**Hispanic participants:**<br>**Intervention:** 341, 341<br>**Control:** 193, 193<br>(their data were combined with other ethnic groups) | 3 studies of 56 weeks<br>1 study of 32 weeks | **Primary:** Efficacy and safety of liraglutide.<br>**Secondary:** Weight and risk factors. | **Yes:** Efficacy and safety were largely similar between Hispanic and non-Hispanic. |
| Sánchez-Rodríguez<sup>28</sup><br>Mexico City<br>2016 | Healthy postmenopausal women or with MetS.<br>Healthy women: Intervention: 52 ± 0.6; Control: 53 ± 0.7 years old.<br>MetS women: Intervention: 52 ± 0.7; Control: 53 ± 0.9 years old. | 100 | **Intervention:** Hormone therapy<br>**Control:** Placebo | **Intervention:** 50, 46<br>**Control:** 50, 45 | 6 months | **Primary:** Oxidative stress. | **Yes:** After 6 months, MetS decreased in the hormone treated group (48%), triglycerides and HDL-c; the controls did not show differences. SS in MSW-HT decreased (3.8 ± 0.3 to 1.7 ± 0.3, p < 0.05) and Oxidative stress was also reduced (44%), this effect was evident since 3 mo. HW-HT with high OS also decreased (40%).<br>In placebo groups there was no change. |
| Mendez-del Villar<sup>42</sup><br>Jalisco<br>2017 | Male and Female<br>Intervention: 41.3 ± 9.7; Control: 54 ± 3.5 years old.<br>BMI: 25-34.9 Kg/m<sup>2</sup><br>T2DM and inadequate glycemic control | 12 | **Intervention:** Metformin + Diacerein<br>**Control:** Metformin | **Intervention:** 6, 6<br>**Control:** 6, 6 | 3 months | **Primary:** Glycemic control. | **Yes:** Significant decrease in fasting glucose, postprandial glucose and A1C. |
| Gonzalez-Heredia<sup>24</sup><br>Jalisco<br>2017 | Male and female<br>Intervention: 49.3 ± 5.7; Control: 51.9 ± 6.4 years old.<br>BMI 25–34.9 Kg/m<sup>2</sup><br>Impaired Glucose Tolerance | 16 | **Intervention:** Linagliptin<br>**Control:** Metformin | **Intervention:** 8, 8<br>**Control:** 8, 8 | 3 months | **Primary:** To assess the effect of linagliptin versus metformin on glycemic variability in patients with IGT. | **Yes:** Group with linagliptin had decrease in glucose levels at 120 min of OGTT.<br>**No:** No significant differences in the AUC, MAGE, SD of glucose, CV of glucose, and MBG between groups. |
| Gonzalez-Ortiz<sup>25</sup><br>Jalisco<br>2017 | Male<br>Intervention: 40.2 ± 7.9; Control: 38.4 ± 6.4 years old.<br>BMI 30-39.9 Kg/m<sup>2</sup> | 18 | **Intervention:** Tadalafil<br>**Control:** Placebo | **Intervention:** 9, 9<br>**Control:** 9, 9 | 28 days | **Primary:** Blood pressure, cholesterol, triglycerides, HDL-c, LDL-c, glucose. | **No:** After the administration of tadalafil there were no significant differences in total insulin secretion first phase of insulin secretion and insulin sensitivity. No significant differences were shown in other measurements. |
| Le Roux<sup>26</sup><br>Multinational<br>2017 | Male and female<br>Intervention: 47.5 ± 11.7; Control: 47.3 ± 11.8 years old. | 2254<br>**Hispanic:** 213 | **Intervention:** Liraglutide<br>**Control:** Placebo | **Intervention:** 1505, 783<br>**Control:** 749, 327 | 40 months (3.3 years) | **Primary:** Evaluate the proportion of individuals with prediabetes who were diagnosed with type 2 diabetes. | **Yes:** Time to onset of diabetes over the 40 months among all randomized individuals was 2.7 times longer with liraglutide than |
| | BMI $\geq 27$ Kg/m<sup>2</sup><br>Dyslipidemia, or<br>hypertension, or<br>both | | | **Hispanic**<br>**participants**<br>**Intervention:** 143<br>**Control:** 70<br>(their data were<br>gathered with other<br>individuals) | | **Secondary:** GLP-1<br>receptor agonist, waist<br>circumference (cm),<br>glycated hemoglobin (%), 2-h plasma glucose during<br>OGTT (mmol/L), Free fatty<br>acids (mmol/L), Blood<br>pressure (mm Hg). | with placebo.<br>Greater weight loss than<br>placebo at month 40: BMI,<br>waist circumference, glycated<br>hemoglobin, fasting glucose,<br>fasting insulin, fasting C-peptide, glucose levels in<br>OGTT, systolic and diastolic<br>blood pressure, and heart<br>rate. |

NIDDM: Non-insulin dependent diabetes mellitus; BMI: Body mass index; T2DM: Type 2 diabetes mellitus; FDA: Food and Drug Administration; A1c: Glycated hemoglobin; HDL: High density lipoprotein; LDL: Low density lipoproteins; VLDL: Very low density lipoprotein; TT: Total testosterone, TL: Free testosterone, SHBG: Sex Hormone Binding Globulin; MetS: Metabolic syndrome; IGT: Impaired glucose tolerance; TG: Triglycerides; ITT: Insulin tolerance test; IL-1 $\beta$ : Interleukin 1 beta; IL-6: Interleukin-6; TNF- $\alpha$ : Tumour Necrosis Factor alpha; WHI: Waist Hip Index; NAFLD: Non-alcoholic fatty liver disease; IR: Insulin resistance; HOMA-IR: Homeostasis model assessment of IR; HOMA B-cells: Homeostatic Model Assessment-beta-cell function; SS: Stress score; MSW-HT: MetS women were assigned to HT (hormone therapy); HW-HT: Healthy-hormone therapy; OS: Oxidative stress; OGTT: Oral glucose tolerance test; AUC: Area under the curve; MAGE: Mean amplitude of glycemic excursion, SD: Standard deviation; CV: Coefficient of variation; MBG: Mean blood glucose; GLP-1: Glucagon-like peptide-1.

**Table 4.** Descriptive characteristics and assessment of Medications.

| Surgical and Alternative Intervention |  |  |  |  |  |  |  |
| --- | --- | --- | --- | --- | --- | --- | --- |
| Author<br>State<br>Year | Participants | Sample size | Intervention<br>implemented/Control | Number of<br>Participants<br>(basal, final) | Treatment<br>duration | Aims/Outcomes | Significance difference<br>between groups |
| Robles-Cervantes<sup>68</sup><br>Jalisco<br>2007 | Female<br>Intervention: 34.0<br>± 3.7; Control:<br>34.6 ± 3.6 years<br>old.<br>BMI: 30-33 Kg/m<sup>2</sup> | 12 | **Intervention:** Liposuction<br>and diet<br>**Control:** Diet | **Intervention:** 6, 6<br>**Control:** 6, 6 | 6 months | **Primary:** Visceral fat, Insulin<br>sensitivity, leptin and tumor<br>necrosis factor alfa.<br>**Secondary:** Glucose,<br>Creatinine, Uric acid, Total<br>cholesterol, HDL cholesterol,<br>Triglycerides. | **Yes:** Leptin correlated with<br>the subcutaneous fat.<br>**No:** no significant difference<br>in insulin sensitivity and did<br>not correlate with<br>subcutaneous fat, leptin, or<br>TNF-alpha. |
| Arceo-Olaiz<sup>67</sup><br>Mexico City<br>2008 | Male and Female<br>Intervention: 36.5<br>± 9.7; Control:<br>37.8 ± 9.6 years<br>old.<br>BMI: 40-55 kg/m2 | 60 | **Intervention:** Laparoscopic<br>roux- en - y gastric bypass<br>(LRYGB)<br>**Control:** banded LRYGB<br>(BLRYGB) | **Intervention:** 30, 30<br>**Control:** 30, 30 | 24 months | **Primary:** Weight loss | **No:** The studied groups did<br>not have significant<br>differences in weight loss at<br>6, 12, and 24 months. The<br>frequency of complications<br>was similar in both groups. |
| García-Vivas<sup>63</sup><br>Durango<br>2014 | Women<br>18-45 years old<br>BMI ≥25 Kg/m<sup>2</sup><br>without known<br>MetS | 138 | **Intervention:** Accupunture<br>**Control:** Sham Accupunture | 138, 99 | 2 months | **Primary:** anthropometric<br>and biochemical | **Yes:** Acupoint catgut<br>embedding therapy +<br>moxibustion produced<br>significant reduction in body<br>weight insulin and HOMA-IR. |
| Alvarado-Reynoso<sup>65</sup><br>Mexico City<br>2019 | Male and Female<br>Intervention: 42.1<br>± 3.2; Control:<br>37.8 ± 3.3 years<br>old.<br>BMI ≥ 25 Kg/m<sup>2</sup> | 45 | **Intervention:** repetitive<br>transcranial magnetic<br>stimulation) rTMS<br>**Control:** sham rTMs | **Intervention:** 22, 18<br>**Control:** 23, 19 | 2 weeks | **Primary:** body weight, food<br>craving, auto perception,<br>general health, depression<br>and anxiety | **Yes:** In the rTMS-treated<br>group reduced body weight,<br>anxiety, and food craving.<br>General health survey<br>domain improved on physical<br>functioning, emotional role,<br>and vitality. The body shape<br>questionnaire improved. |
| Hernandez-Lepe<sup>64</sup><br>Chihuahua<br>2019 | Male<br>25 ± 5 years old<br>BMI >25 Kg/m<sup>2</sup><br>Sedentary | 52 | **Intervention:** spirulina<br>maxima<br>**Control:** placebo | **Intervention:** 26, 26<br>**Control:** 26, 26 | 3 months | **Primary:** plasma lipid profile<br>and antioxidant capacity | **Yes:** BMI, total cholesterol,<br>triglycerides and LDL-C<br>decreased. HDL-C increased<br>in all treatment groups.<br>Participants with known<br>dyslipidemia had higher<br>response. |

### Nutritional/Behaviour interventions

The comparisons between active nutrition/behavior interventions with placebo showed improvement for BMI [Cohen-d, 95% CI, Figure 3] 0.2 (0.01, 0.38), waist circumference 0.27 (0.01, 0.53, Figure 4), triglycerides 0.34 (-0.02, 0.71; Figure 5), and systolic blood pressure 0.21 (-0.07, 0.49, Figure 6). The lowest heterogeneity was for BMI (I^2= 41%) and the highest for triglycerides (I^2= 88%). Only one intervention with physical activity showed an effect on BMI (Cohen-d of 0.3), increase on HDL-c (Cohen-d 0.16), but with wide confidence intervals. Most of these studies excluded T2D individuals, therefore the glucose levels did not show difference between compared groups.

**Figure 4.**
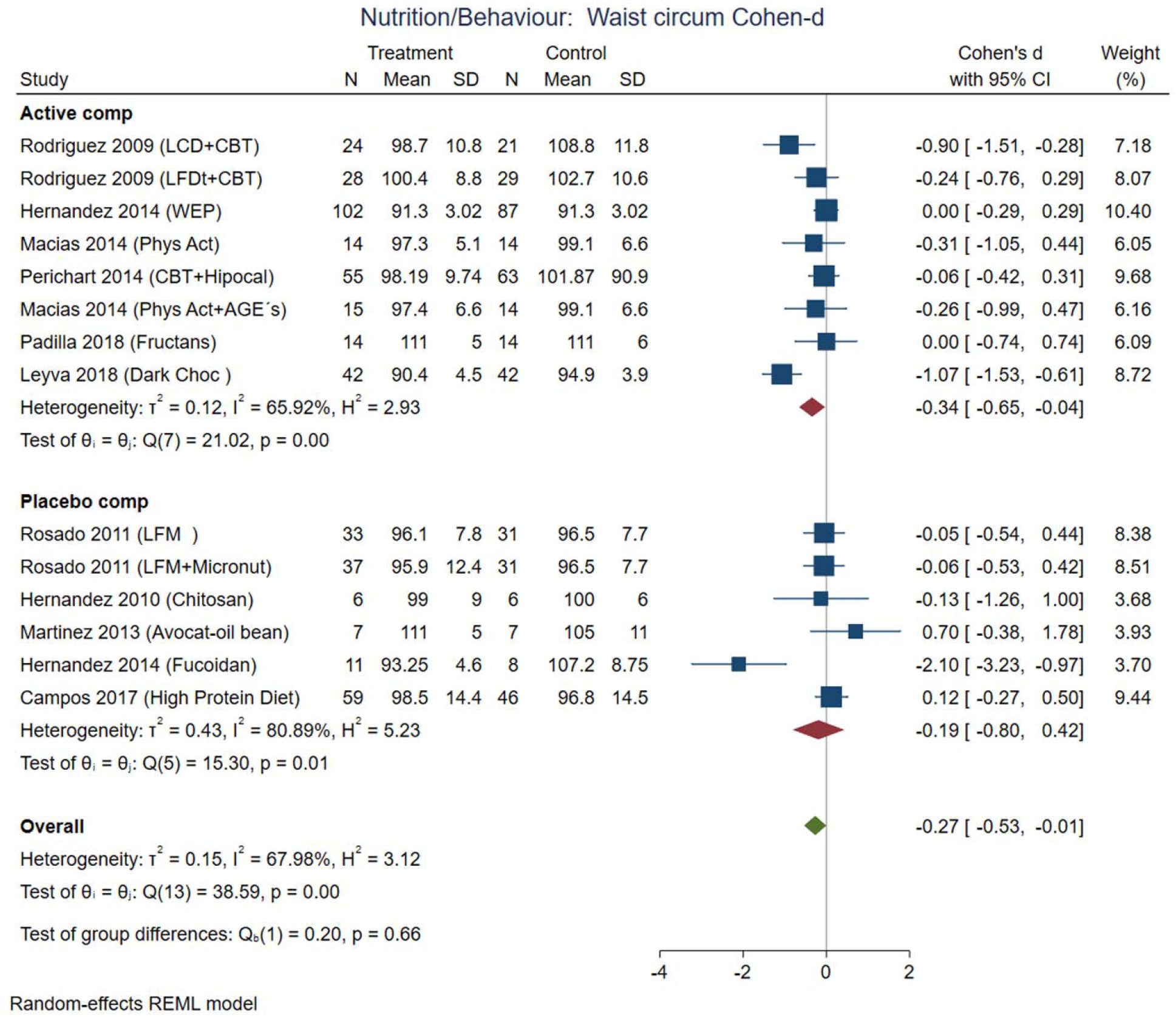
Pooled analysis of weighted size of effect by Cohen-d in waist circumference with nutritional and behavior interventions. The analysis was stratified by placebo or active comparator. LFDT: Low fat diet, LCD: Low carbohydrate diet, CBT: Cognitive-behavior therapy, Phys Act: Physical activity, Dark Choc: Dark chocolate, AGE: Advance glycation end-product, Hipocal: Hypocaloric diet, LFM: Low fat milk, Micronut: Micronutrients, PMR: Partial meal replacement. REML: Restricted maximum likelihood.

**Figure 5.**
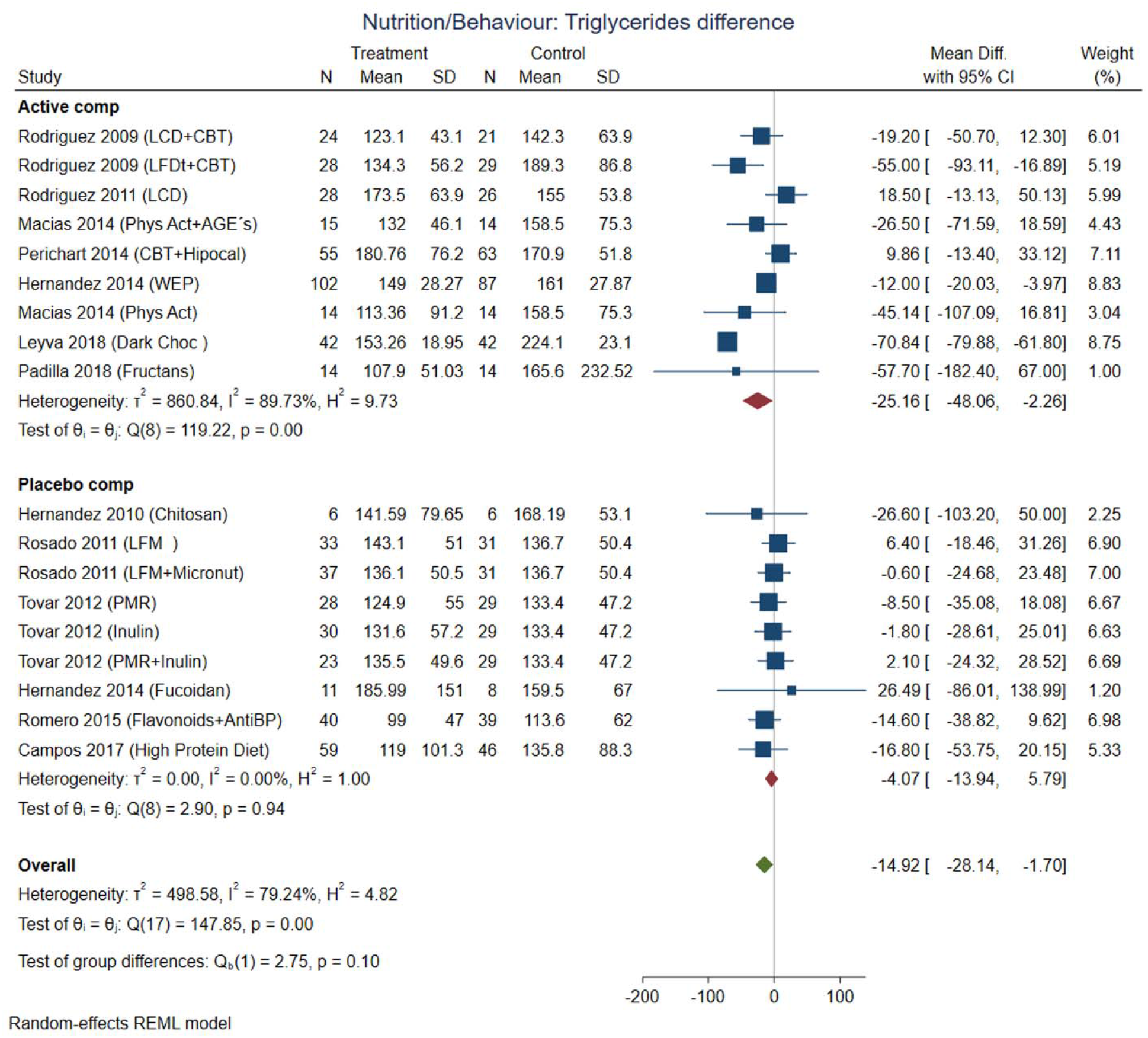
Pooled analysis of weighted size of effect by Cohen-d in triglycerides serum concentration with nutritional and behavior interventions. The analysis was stratified by placebo or active comparator. LFDT: Low fat diet, LCD: Low carbohydrate diet, CBT: Cognitive-behavior therapy, Phys Act: Physical activity, Dark Choc: Dark chocolate, AGE: Advance glycation end-product, Hipocal: Hypocaloric diet, WEP: Water and Education Provision, LFM: Low fat milk, Micronut: Micronutrients, PMR: Partial meal replacement, AntiBP: Antihypertensive medication. REML: Restricted maximum likelihood.

**Figure 6.**
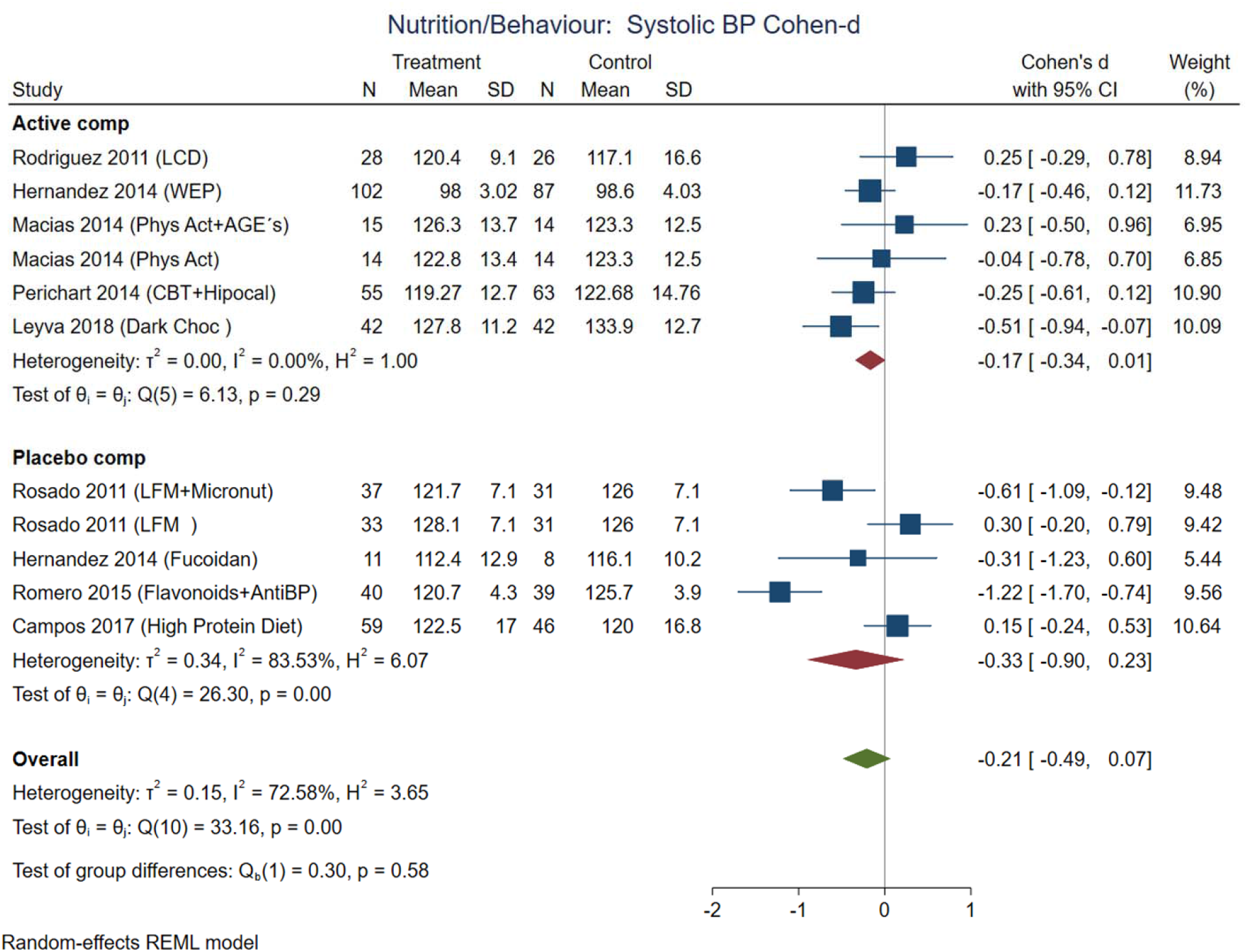
Pooled analysis of weighted size of effect by Cohen-d in systolic blood pressure with nutritional and behavior interventions. The analysis was stratified by placebo or active comparator. LCD: Low carbohydrate diet, CBT: Cognitive-behavior therapy, Phys Act: Physical activity, Dark Choc: Dark chocolate, AGE: Advance glycation end-product, Hipocal: Hypocaloric diet, WEP: Water and Education Provision, LFM: Low fat milk, Micronut: Micronutrients, AntiBP: Antihypertensive medication. REML: Restricted maximum likelihood.

The addition of adding CBT (goal setting, problem-solving, and stimulus control) to either a low-fat diet (21% fat, less than 10% saturated fat, 25% protein, 54% carbohydrates), or a low-carbohydrate diet (27% protein, 28% fat, 45% carbohydrate) produced significantly greater short-term weight loss compared with diet alone. The use of antioxidants with flavonoids contained in dark chocolate showed favorable changes in biochemical parameters (total cholesterol, triglycerides, and LDL-cholesterol level in blood) and anthropometrical parameters (waist circumference) the pooled analysis with Cohen-d supported additionally loss of BMI and decrease in systolic blood pressure (Figure 6).

Finally, the avoid of sugar-sweetened beverages (SSB) by water substitution showed positive effect on plasma triglycerides, and systolic blood pressure.

### Drug treatments and T2D status

Drug treatment analyzed studies which involved participants with T2D. The size of effect showed improvement on BMI (Figure 7), waist circumference (Figure 8) and glucose (Figure 9) for non-T2D individuals compared with patients with T2D. The BMI loss in T2D group had a Cohen-d of 0.24 (0.13, 0.66) compared with non-T2D loss of 0.53 (0.27, 0.80); the waist circumference was Cohen-d 0.22 (0.72, 1.16) compared with non-T2D 0.55 (0.03, 1.07); diastolic blood pressure 0.18 (0.3, 1.42) vs 0.87 (0.33, 2.06), respectively. As expected, the treatment had large effect on glucose lowering for treated T2D individuals compared with non-T2D (Cohen-d 0.7 compared with 0.26, respectively).

**Figure 7.**
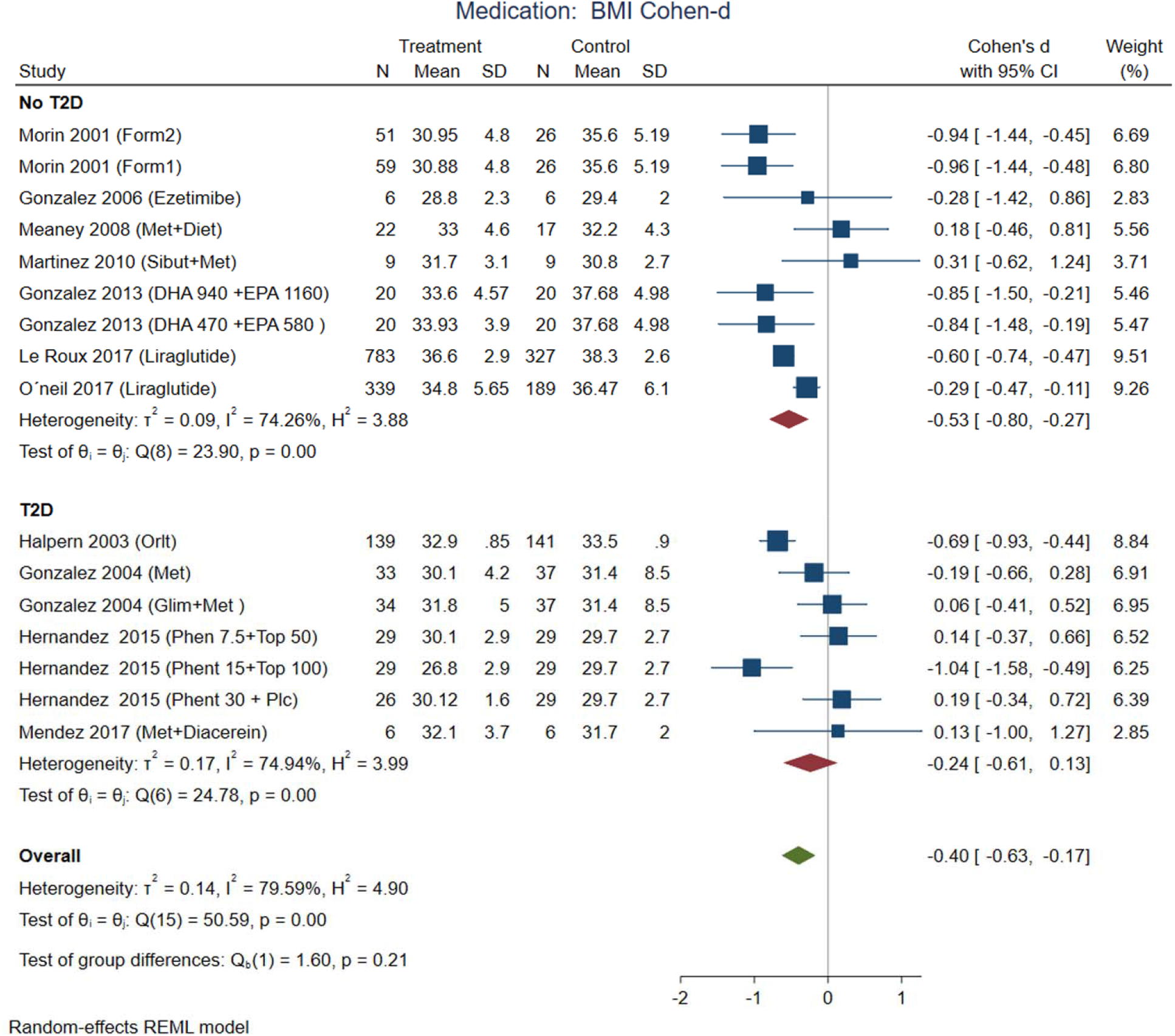
Pooled analysis of weighted size of effect by Cohen-d in BMI loss with drug (medication) treatment. The analysis was stratified by T2D status. The Form1 and Form2 are described in the text, they are not approved by FDA. Met: Merformin, Sibut: Sibutramine, DHA: Docosahexaenoic acid, EPA: Eicosapentanoic acid, Orlit: Orlistat, Glim: Glimepiride, Phent: Phentermine, Top: Topiramate. REML: Restricted maximum likelihood.

**Figure 8.**
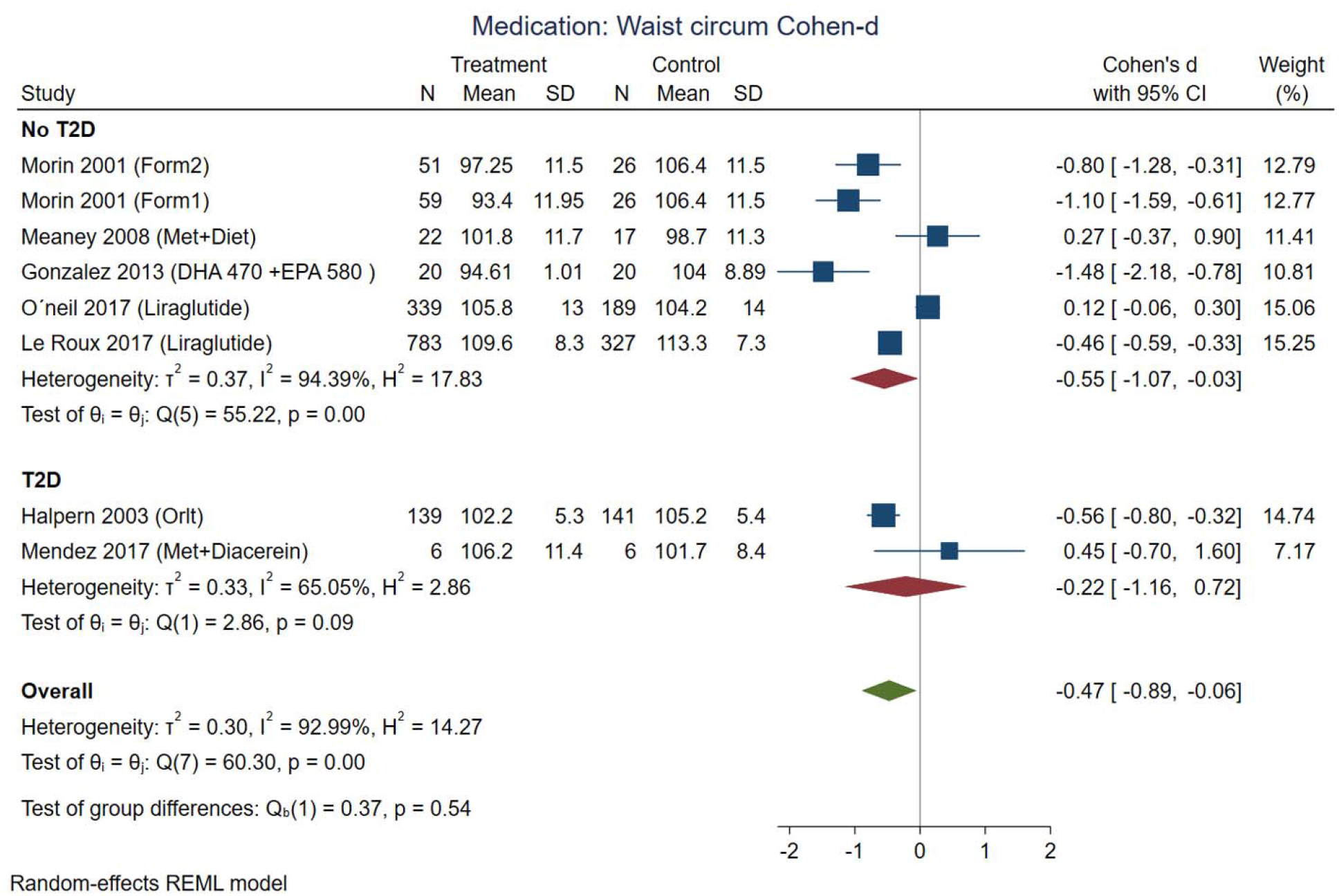
Pooled analysis of weighted size of effect by Cohen-d in waist circumference with drug (medication) treatment. The analysis was stratified by T2D status. The Form1 and Form2 are described in the text, they are not approved by FDA. Met: Merformin, Sibut: Sibutramine, DHA: Docosahexaenoic acid, EPA: Eicosapentanoic acid, Orlit: Orlistat, Glim: Glimepiride, Phent: Phentermine, Top: Topiramate. REML: Restricted maximum likelihood.

**Figure 9.**
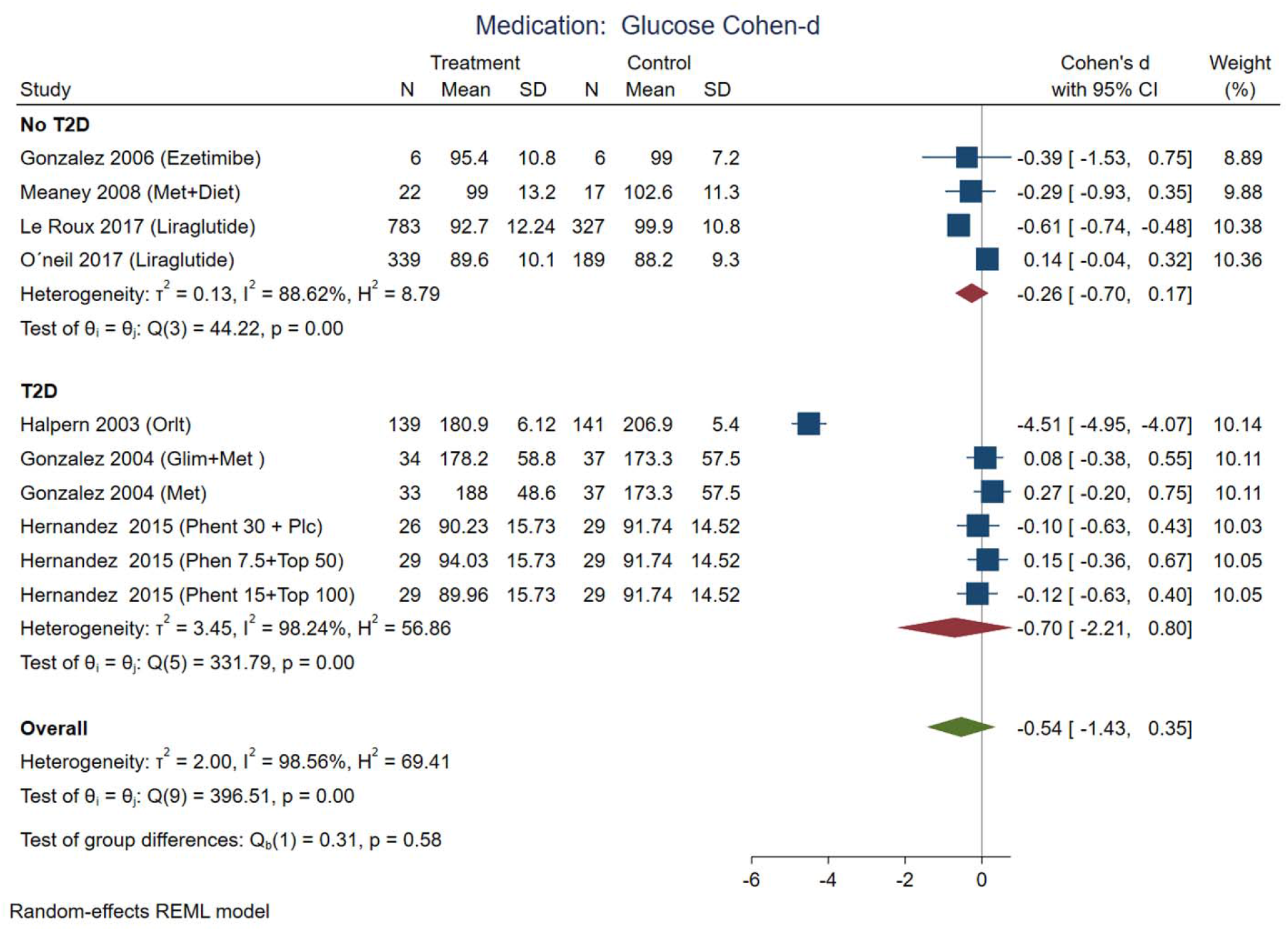
Pooled analysis of weighted size of effect by Cohen-d in glucose serum concentration with drug (medication) treatment. The analysis was stratified by T2D status. Met: Merformin, Orlit: Orlistat, Glim: Glimepiride, Phent: Phentermine, Top: Topiramate. REML: Restricted maximum likelihood.

Some medications used in Mexico had a large effect on weight reduction (Figures 7 and 8) in participants without T2D (Cohen-d about 0.9). For instance, the use of DHA (docosahexanoic acid) 470 or 940 mg combined with EPA (eicosapentanoic acid) 580 or 1160 mg, compared with placebo, and the use of two different formulations (Formula 1: d-norpseudoephedrine 50 mg, triiodothyronine 75 ug, diazepam 5 mg, atropine 0.36 mg, aloin 16.2 mg; and formula 2: d-norpseudoephedrine 50 mg, atropine 0.36 mg, aloin 16.2 mg.) for 6 months compared with placebo. These medications are not approved for treatment of obesity by FDA, and the formulations are not legally available for purchase in the US, however, reports in US found thyroid intoxication (70). The effect of liraglutide was between 0.3 to 0.6 including participants from international samples. Participants with T2D showed the use of phentermine 15 mg and topiramate 100 mg had higher effect compared with phentermine 7.5 mg and placebo. There was no replication for any of these studies, the Egger test on a random model showed no small study effects on BMI for 18 intervention on nutrition/behavior (p=0.43), nor for 19 intervention on medication (p=0.22).

It is interesting that systolic blood pressure was modified by non-pharmacological treatments, meanwhile, diastolic blood pressure was modified in non-T2D participants treated with medications (Figure 10). From the five analyzed studies with medications, three of them included patients with hypertension. The prevalence of hypertension was between 24 to 42%. The blood pressure decreases with weight loss, the Trial of Hypertension Prevention had a weight loss intervention arm, resulting in reduction of both, systolic and diastolic, measurements (71).

**Figure 10.**
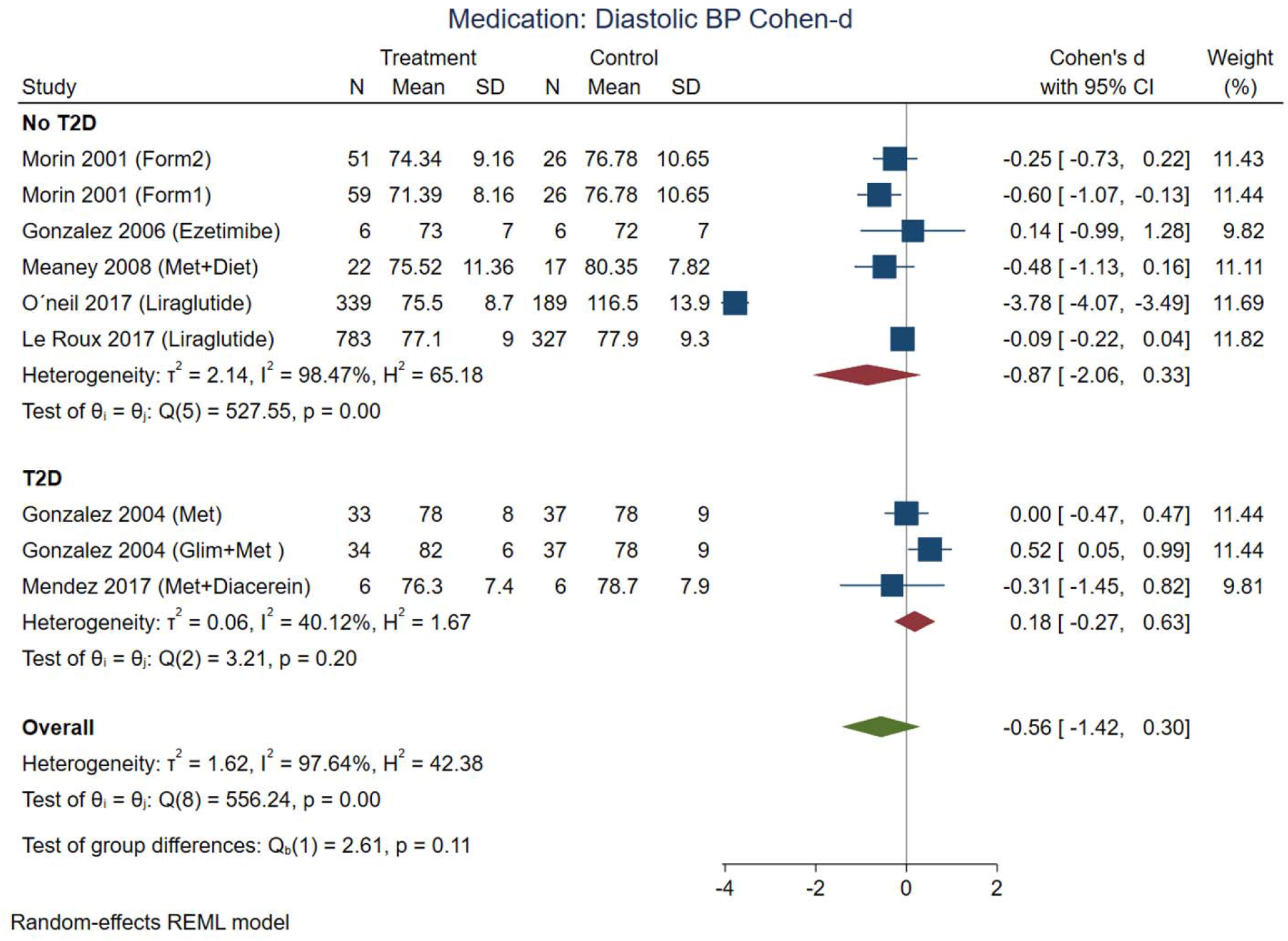
Pooled analysis of weighted size of effect by Cohen-d in diastolic blood pressure with drug (medication) treatment. The analysis was stratified by T2D status. The Form1 and Form2 are described in the text, they are not approved by FDA. Met: Merformin, Sibut: Sibutramine, Orlit: Orlistat, Glim: Glimepiride, Phent: Phentermine, Top: Topiramate. REML: Restricted maximum likelihood.

### Study heterogeneity sources

The meta regression analyzed the mean age, BMI, months of treatment, comparison with placebo and geographical location measured by latitude. Those confounders that reached statistical significance for HDL-c serum levels were the duration of the intervention [b=1.07 (se 0.49) p=0.03] and the comparison vs placebo [b=4.4 (se 2.1) p=0.04]. The triglyceride serum levels showed effects from the mean age of the study [b=1.3 (se 0.59) p=0.03] and the geographic location [b= -2.7 (se 1.3) p=0.04]. However, the geographic location was close related with the type of intervention, for example, studies located close to the U.S.-Mexico border used physical activity interventions; meanwhile the South regions used nutritional supplements. The diastolic blood pressure was modified by the BMI [b=-0.49 (se 0.27) p=0.067] and geographical location [b= - 0.9 (se 0.44) p=0.04], however these variables were influenced by the treatment with liraglutide. [Supplemental Figures 1 and 2]

### Network meta-analysis

A network meta-analysis of drug treatments and T2D status was performed for BMI, diastolic blood pressure (DBP), and glucose. The network meta-analysis included direct comparisons constructed with connections between treatments, and indirect comparisons using all possible connections between treatments. All networks had the principles of coherence, transitivity, and consistency. This analysis was not feasible for nutritional/behavior interventions due to the design and the small number of studies. Figure 11 illustrates two networks for studies with T2D patients, examining the efficacy of pharmacological interventions on the studied variables, one network for each comparison between treatments and placebo (Supplemental figure 3 panel A) and with metformin (Supplemental figure 3 panel B). These network diagrams provide a graphical representation of how each intervention connects to any other direct comparisons. Tables 5 (matrix A and B) and Supplemental Tables 1 and 2 detail the complete matrix of results, in which the comparative effects between drugs are shown in terms of differences in means standardized.

**Table 5.** Network meta-analysis results matrix.

| **Matrix A: Drugs (Non-diabetic participants)** |  |  |  |  |  |
| --- | --- | --- | --- | --- | --- |
| **BMI (Reference: placebo)** |  |  |  |  |  |
| **DHA 470 + EPA 580** | 0.072 (-0.672, 0.815) | 0.143 (-0.852, 1.138) | 0.129 (-0.872, 1.130) | -0.365 (-1.192, 0.462) | -0.816 (-1.582, -0.049) |
| -0.072 (-0.815, 0.672) | **DHA 940 + EPA 1160** | 0.071 (-0.927, 1.070) | 0.057 (-0.947, 1.061) | -0.436 (-1.267, 0.394) | -0.888 (-1.658, -0.117) |
| -0.143 (-1.138, 0.852) | -0.071 (-1.070, 0.927) | **Form1** | -0.014 (-0.570, 0.541) | -0.508 (-1.214, 0.198) | -0.959 (-1.593, -0.324) |
| -0.129 (-1.130, 0.872) | -0.057 (-1.061, 0.947) | 0.014 (-0.541, 0.570) | **Form2** | -0.493 (-1.208, 0.221) | -0.944 (-1.588, -0.301) |
| 0.365 (-0.462, 1.192) | 0.436 (-0.394, 1.267) | 0.508 (-0.198, 1.214) | 0.493 (-0.221, 1.208) | **Liraglutide** | -0.451 (-0.761, -0.141) |
| 0.816 ( 0.049, 1.582) | 0.888 ( 0.117, 1.658) | 0.959 ( 0.324, 1.593) | 0.944 ( 0.301, 1.588) | 0.451 ( 0.141, 0.761) | **Plc** |
| **Matrix B: Drugs (Diabetic participants)** |  |  |  |  |  |
| **BMI (Reference: metformin)** |  |  |  |  |  |
| **Diac+Met** | -0.080 (-1.307, 1.147) | -0.143 (-1.374, 1.088) | -0.774 (-2.026, 0.478) | 0.124 (-1.009, 1.257) |  |
| 0.080 (-1.147, 1.307) | **Glim** | -0.063 (-0.529, 0.403) | -0.694 (-1.404, 0.017) | 0.204 (-0.266, 0.675) |  |
| 0.143 (-1.088, 1.374) | 0.063 (-0.403, 0.529) | **Glim+Met** | -0.631 (-1.349, 0.087) | 0.267 (-0.214, 0.749) |  |
| 0.774 (-0.478, 2.026) | 0.694 (-0.017, 1.404) | 0.631 (-0.087, 1.349) | **Insulin** | 0.898 ( 0.366, 1.431) |  |
| -0.124 (-1.257, 1.009) | -0.204 (-0.675, 0.266) | -0.267 (-0.749, 0.214) | -0.898 (-1.431, -0.366) | **Met** |  |

Matrix A represent the estimates of the effect of treatments (standardized mean differences and 95%CI) relative to placebo. The dosage of DHA and EPA are in mg per day. Form1: D-norpseudoephedrine 50 mg, triiodothyronine 75 ug, diazepam 5 mg, atropine 0.36 mg, aloin 16.2 mg. Form2: D-norpseudoephedrine 50 mg, atropine 0.36 mg, aloin 16.2 mg. These two formulations are not approved by FDA. Plc: Placebo. Matrix B shows estimates of the effect of treatments (standardized mean differences) relative to metformin in patients with diabetes. Diac: Diacerin; Gim: Glimepiride; Met: Metformin.

The contrast matrix between pharmacological treatments with placebo showed a decrease on BMI by any pharmacological intervention, for instance, DHA 470 mg + EPA 580 mg was 0.816 (CI: 0.049, 1.582); DHA 940 mg + EPA 1160 mg: 0.888 (CI: 0.117, 1.658); Formulation 1: 0.959 (CI: 0.324, 1.593); Formulation 2: 0.944 (CI: 0.301, 1.588); Liraglutide: 0.451 (CI: 0.141, 0.761). On the other hand, the status of T2D consistently supported metformin alone and in combinations were the most effective intervention for reducing BMI compared to insulin: -0.898 (CI: -1.431, -0.366). Regarding glucose, the intervention with insulin was more effective in reducing serum glucose levels compared to metformin: - 1.506 (CI: -2.084, -0.928); Glimepiride + metformin: -1.332 (CI: -2.083, -0.581) and Glimepiride: -1.332 (CI: -2.077, -0.587). In summary, the interventions with the greatest contribution to the reduction of DBP were metformin: -0.507 (CI: -0.994, -0.020) compared to Glimepiride + metformin, and Glimepiride: - 0.507 (-0.980, -0.033) compared to Glimepiride + metformin. The monotherapy interventions have better effectiveness in DBP compared to double therapies.

## Discussion

This systematic review and meta-analysis summarize the existing evidence of weight loss as primary or secondary aims in adult population. Our analysis was limited to randomized clinical studies conducted in Mexico or from international multicentric studies with Mexican participants involving nutrition, behavior, medication, or alternative medicine interventions. Some interventions of interest were compared with another active strategy (medication, behavior, physical activity or any different than placebo), this strategy can blunt the size of effect of the intervention, because the effect of active comparators in metabolic and anthropometric variables. We found that all studied interventions were better than placebo, or better than the selected comparator, and many of the published papers made individual paired contrasts between final and basal values. However, we decided to contrast treatments and reported the size of effects by metabolic syndrome component. With this strategy we had the advantage of computing the effect size over a maneuver the researchers considered the best comparator. The results should be interpreted considering these control groups defined by the researchers.

### Interventions

The 55 analyzed interventions (from 45 studies) were categorized in nutritional/behavior with a total sample of 1,407 participants; drug interventions in Mexico included 1,134; and multinational interventions were additional 1,307 participants (Hispanics); surgical procedures were 72 and alternative treatments 235 individuals. We obtained a total of 4,155 participants from these trials.

The nutritional/behavior strategies included supplemental, flavonoids, manipulation of macronutrient content diets (low fat, low carb, high protein) with caloric restriction, water consumption and physical activities. Cognitive behavioral therapy (CBT) combined with low calorie diet showed beneficial effects on BMI and waist circumference; and combined with low fat diet deceased glucose, triglycerides. A cardioprotective structured hypocaloric diet is more effective than the CBT approach in reducing metabolic syndrome (59). Daily flavonoid-rich chocolate (70% cocoa) intake improves fasting plasma glucose levels and insulin resistance parameter (HOMA-IR) and the lipid and glucose metabolism (46). The physical activity showed benefic but small and non-significant effects for the analyzed variables, due to the lack of enough sample size. Other systematic reviews focused on physical activity showed Hispanics had less leisure-time compared with other groups in the U.S., the most common activity was walking, but the most significant results were those with moderate to vigorous physical activity (72). It will be crucial to increase legislative policies to build environments that increase available opportunities for physical activities, particularly for this fast-growing population group.

Adherence to diet and exercise programs (45 min-60 min/d, 5 days per week) are part of the nutritional/behavioral interventions. Other studies reporting that water consumption habit (2–3 L/day) and partially decreasing sugar-sweetened beverage (SSB) intake of at least 250 kcal/d, with nutritional counseling was effective in increasing water intake (69), and additionally reduces cardiometabolic risks of drinking or eating less sugar in the diet promoting health benefits, although we found positive effect on plasma triglycerides and systolic blood pressure in our analysis, perhaps a consequence of the reduction of the SSB consumption.

The drug treatment with groups of participants with T2D, showed small effect size on improvement on BMI, waist circumference and triglycerides compared with larger effects for non-T2D. The orlistat group in T2D showed weight loss (BMI and waist circumference) lower level of glucose, triglycerides, and systolic blood pressure. Comparing these findings with other studies made in Mexican Americans living in the border shows the difficulty of losing weight with programs on self-management education, but the HbA1c improved (73). No medication will overcome unhealthy lifestyles.

Medication showed a larger size of effects on BMI for combined formulations like orlistat, phentermine with topiramate, both approved by regulatory agencies. Other formulations like the combination of triiodothyronine with phentermine (non-approved by FDA but approved by COFEPRIS – Federal Committee for Protection from Sanitary Risks), and combination of DHA and EPA showed effect on BMI. The authors of the formulations did not show the result on serum glucose neither reported any adverse effect. There was no replication for any of these treatments. We found a couple of sibutramine trials. This is a retired medication because the cardiovascular risk was greater than the benefits (74), specially for the difficulty to identify patients with silent cardiovascular disease (75).

Surgical intervention is the most effective treatment for patients with morbid obesity.(76) The percentage of body weight loss with this intervention ranges between 33 and 77% in a period of 24 months, thus demonstrating its effectiveness (77, 78). However, in our surgical papers, no significant differences were found in the percentage of weight loss, this due to the fact that both the intervention group and the control group had equivalent surgeries (79). One of the studies compared banded versus unbanded laparoscopic roux-en-Y gastric bypass and follow up weight changes for 24 months,(67) in a second analysis no differences were found between these procedures after five years of follow up (66).

### Risk of bias

In general, many of the studied interventions are challenging to blind for obvious reasons. For example, a comparison of nutritional interventions vs. exercise or CBT cannot be blind. However, there is a possibility to blind the evaluators, but no studies explicitly describe this strategy. We found that heterogeneity of the results were partially attributable to basal differences between contrasting groups, for example in the study of Rosado et al.,(51) the diastolic and systolic blood pressure were significantly different between the studied low fat milk groups compared with controls. Some surgical studies for weight loss made in the Instituto de Nutricion Salvador Zubiran in Mexico City blinded the abdominal wall for patients and evaluators when they compared the open abdominal approach versus the laparoscopic method. The risk of bias can be lessened but still can compromise the results of the studies. The difficulty in addressing nutritional or behavioral interventions is manifest in studies analyzing racial/ethnic disparities. Multilevel church-based interventions considering socio-ecological influence showed a greater impact if they consider program interventions tailored to specific communities.

### Limitations

The most important limitations are the lack of replication studies with the same medications, and the small sample size for most of the studies. There was wide variety in the criteria for selection of the participants (i.e.: some studies had too specific eligibility criteria for sex, age and BMI compared with other studies with wide range of options), and, despite similar genetic background, the participants live in sites embedded in cultural diversity (i.e.: Mexico City’s environment problems differ from those in States close to the U.S.-Mexico border). We address a broad question regarding the metabolic syndrome components with important heterogeneity of the studies. We addressed this problem using meta-regression to statistically weight the main confounders across studies and the use of a network meta-analysis to compute the magnitude of contrasts between treatment effects. Due to these limitations the obtention of unstable coefficients is possible, therefore, the analysis should be repeated in the future with a greater number of studies.

The small sample sizes from many of the included studies resulted in low statistical power for contrasting between treatment, and the lack of replication studies increased the standard error for the analysis. The new medications approved by FDA have been tested scarcely in Mexican population. About 44% of the studies were performed in the limit time of placebo effects (about 12 weeks), but those with more time showed effects on the HDL cholesterol levels.

Future new and replication studies should consider larger periods for treatments to reduce placebo effects. Future reviews and meta-analysis should analyze anti-obesity interventions in children and adolescents as well old age populations. These suggestions agree with the Healthy People 2030 recommendation on study effective strategies to diminish obesity in children and adolescents (80).

The Mexican states conducted research in anti-obesity interventions were ten from 32 states. The U.S.- Mexico border has sister states: California-Baja California, Arizona-Sonora, New Mexico-Chihuahua, Texas with Chihuahua, Coahuila, Nuevo Leon and Tamaulipas. The Binational initiative should improve the collaborative studies in the U.S.-Mexico border to address interventions in this population. The programs from this initiative addresses environmental protection, communication committees in particular communities (81). The U.S.-Mexico Border Health Commission has agreements with the Secretary of Health from both countries, and this agency supports initiatives in health security (82). The programs include prevention and wellness using guidelines for eating healthy, physical activity, and of drug misuse and abuse prevention.

## Conclusions

Since 1996, anti-obesity interventions have been conducted in Mexico in randomized controlled clinical studies, mainly focused on pharmaceutical, nutritional, or physical activity interventions. Adult participants included in these studies were predominantly from the central and northern Mexican states, with a clear absence from the costal and southern states. Anti-obesity studies in the Mexican population include small samples and reduced time for interventions. A strategy to improve the statistical power for the studies is to conduct multicentric studies, and a compromise for the State or private industries to provide sufficient finantial resources.

A national web of research is feasible for answering relevant questions regarding anti-obesity interventions and its metabolic consequences. It is clear that not all metabolic syndrome components have the same response to the intervention. The inclusion of Mexican Americans and Mexican immigrants living in the U.S. would be desirable to clarify the importance of different techniques to tackle this problem.

## Data Availability

All data are available from well-knwon repositories.

https://pubmed.ncbi.nlm.nih.gov/

https://apps.webofknowledge.com/

https://www.scopus.com/home.uri

## Acknowledgement

We thanks Dr.Victoria Valles from the National Institute of Medical Sciences and Nutrition Dr. Salvador Zubiran (INCMNSZ) for the share of invaluable experience about the ENEC-93. Last, we would like to pay our gratitude and our respects to Dr. Gonzalez-Barranco. He was a pioneer in obesity research since 1970s, Dr. Gonzalez-Barranco passed away in December of 2020.

---

Supplemental Figure 1. Meta regression of medication mean difference effects on HDL-C (upper panel) and triglycerides (lower panel) concentrations, adjusted by mean age, BMI, duration of treatment (months), geographical latitude and use of placebo or active comparator. The grey zone represents the 95%CI of the regression. Liraglutide was used in the highest obesity and geographical sites at North. The Form1 and Form2 are described in the text, they are not approved by FDA. Met: Merformin, Sibut: Sibutramine, Orlit: Orlistat, Glim: Glimepiride, Phent: Phentermine, Top: Topiramate.

Supplemental Figure 2. Meta regression of medication mean difference effects on diastolic blood pressure adjusted by mean age, BMI, duration of treatment (months), geographical latitude and use of placebo or active comparator. The upper panel shows the effect of BMI and the lower panel the geographical location. The grey zone represents the 95%CI of the regression. Liraglutide was used in the highest obesity and geographical sites at North. The Form1 and Form2 are described in the text, they are not approved by FDA. Met: Merformin, Sibut: Sibutramine, Orlit: Orlistat, Glim: Glimepiride, Phent: Phentermine, Top: Topiramate.

Supplemental figure 3. Network meta-analysis of the studies examining the efficacy of drug treatments in patients with obesity in (A) BMI in non-diabetic patients compared to placebo, (B) BMI in patients with diabetes compared to metformin. The colors of edges and nodes refer to the risk of bias: low (green), moderate (yellow), and high (red). The dosage of DHA and EPA are in mg per day. Met: Metformin. Diac+Met: Diacerin + Metformin. The Form1 and Form2 are described in the text, they are not approved by FDA. Plc: Placebo.

