## Supplemental Figures for "Looking for crumbs in the obesity forest: anti-obesity interventions in the Mexican population. History, and systematic review with Meta-Analysis"

Sup
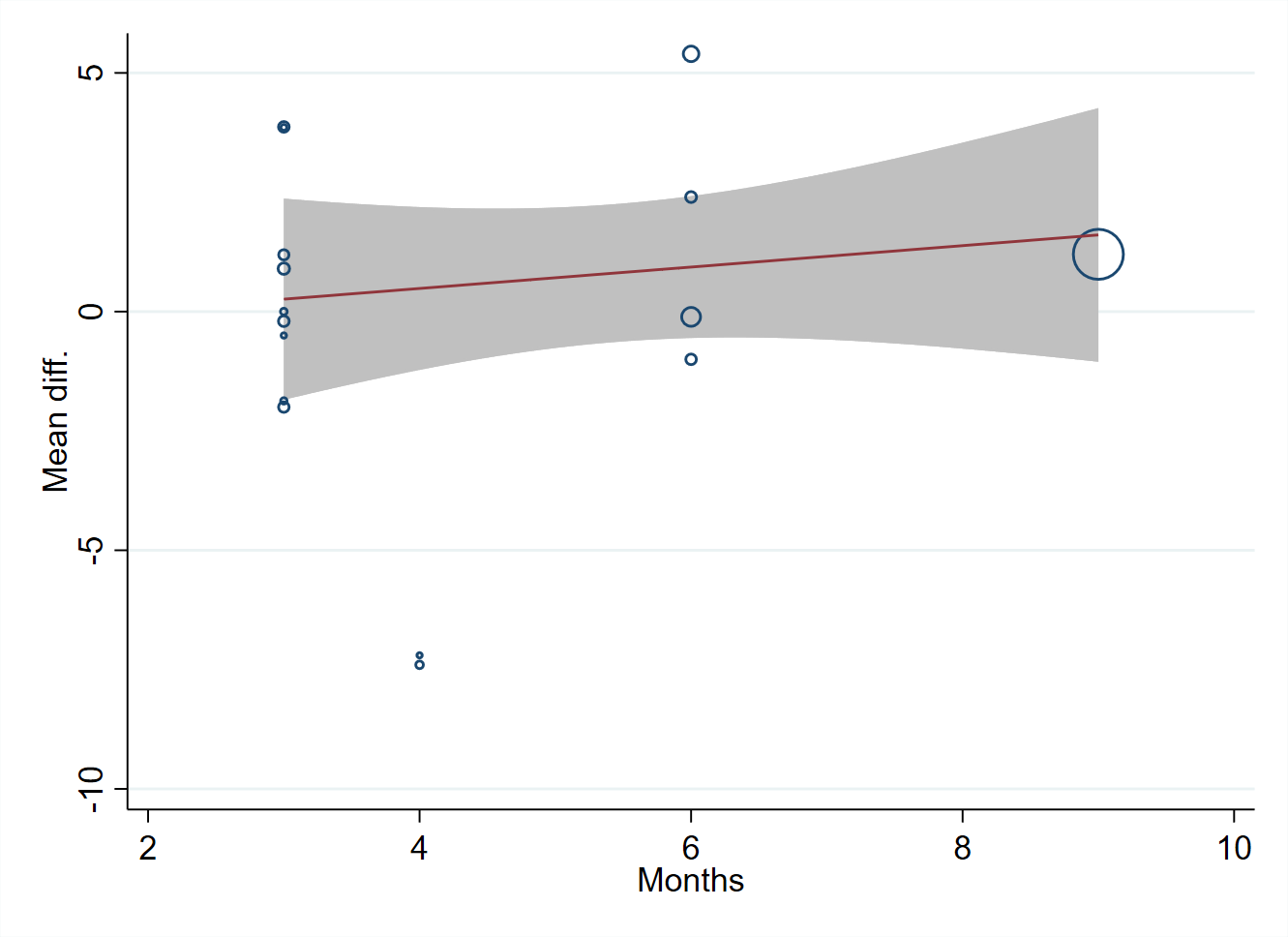

Supplemental Figure 1, panel A.

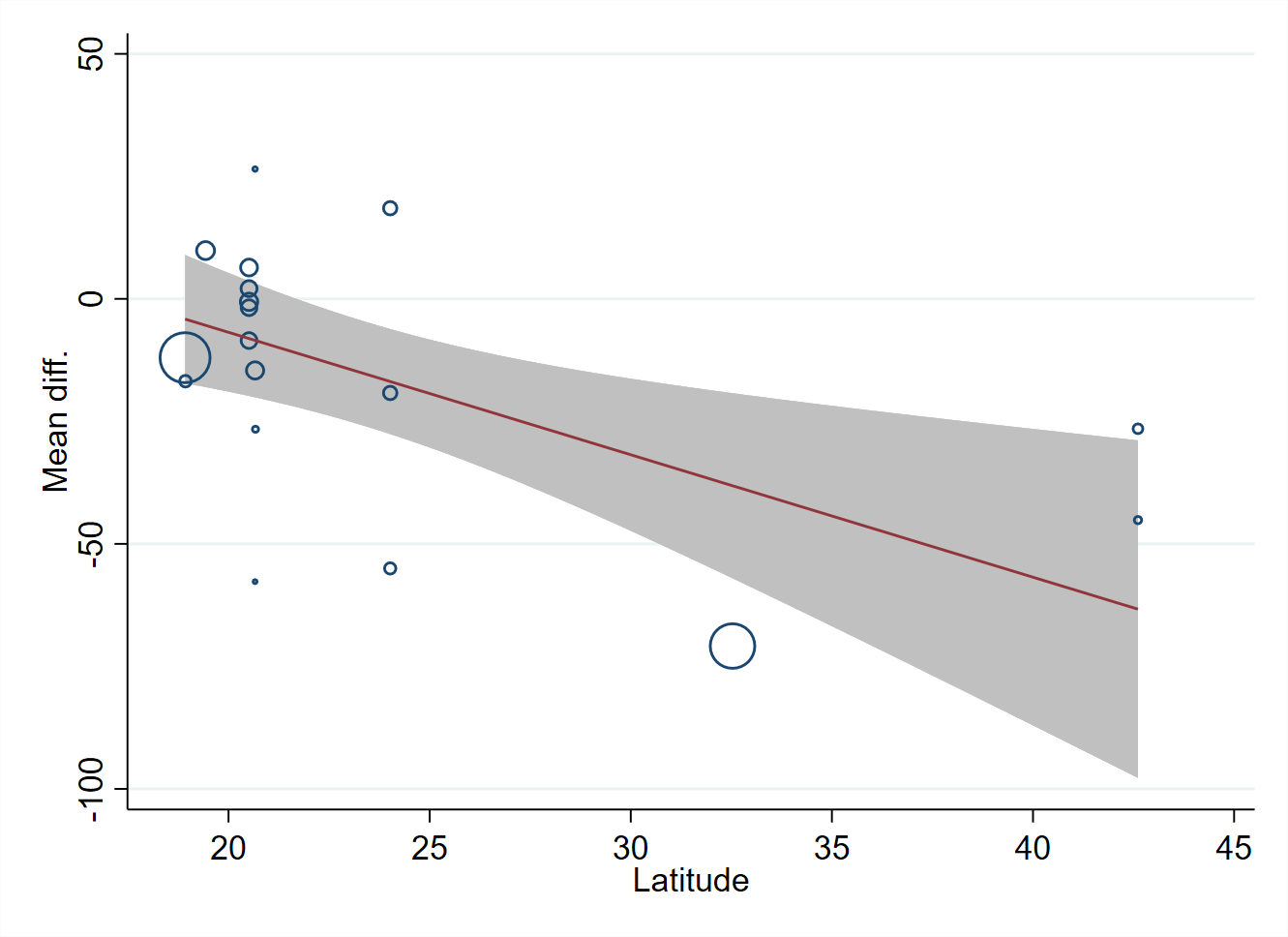

Supplemental Figure 1, panel B.

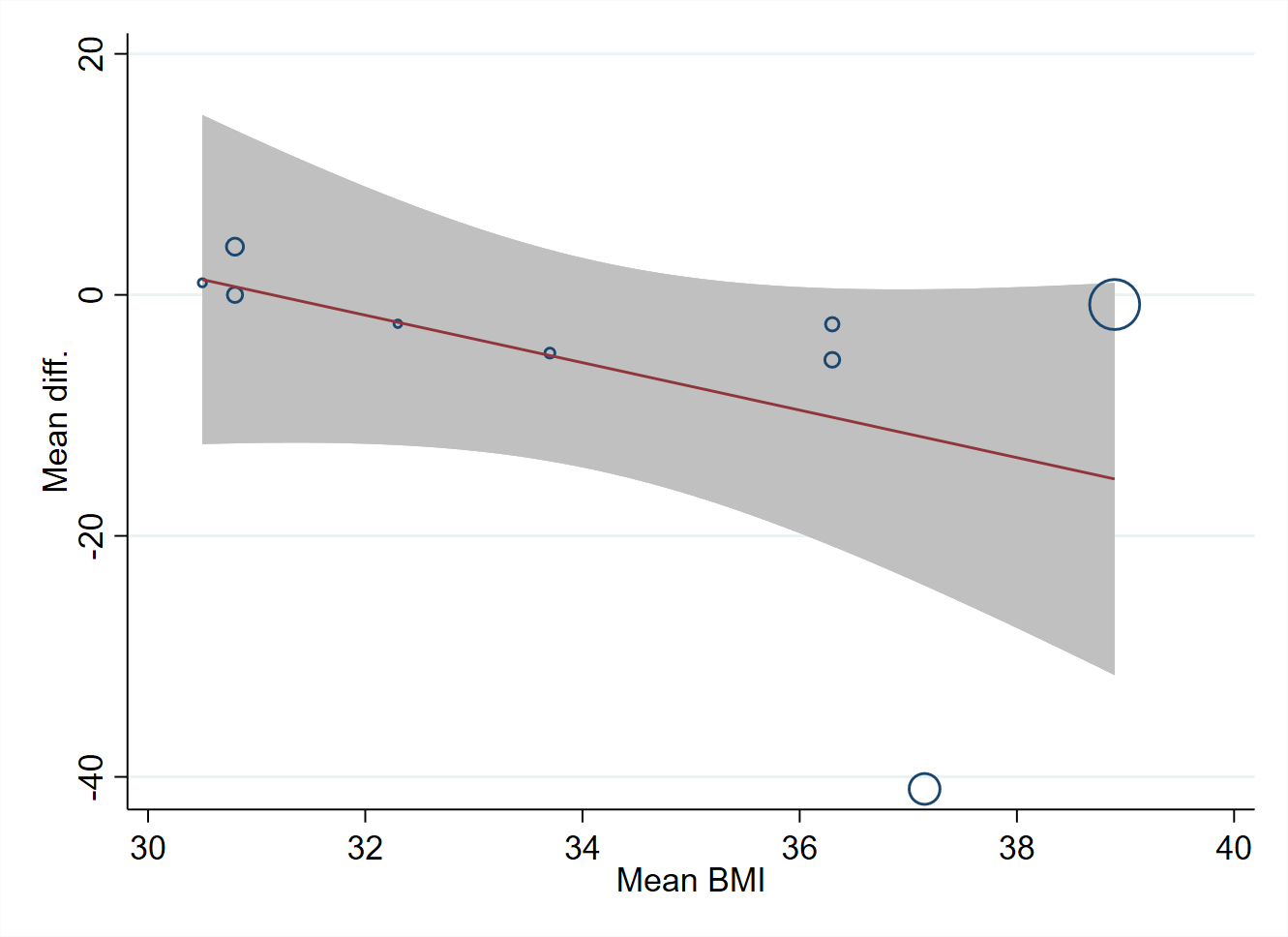

Supplemental Figure 2, panel A.

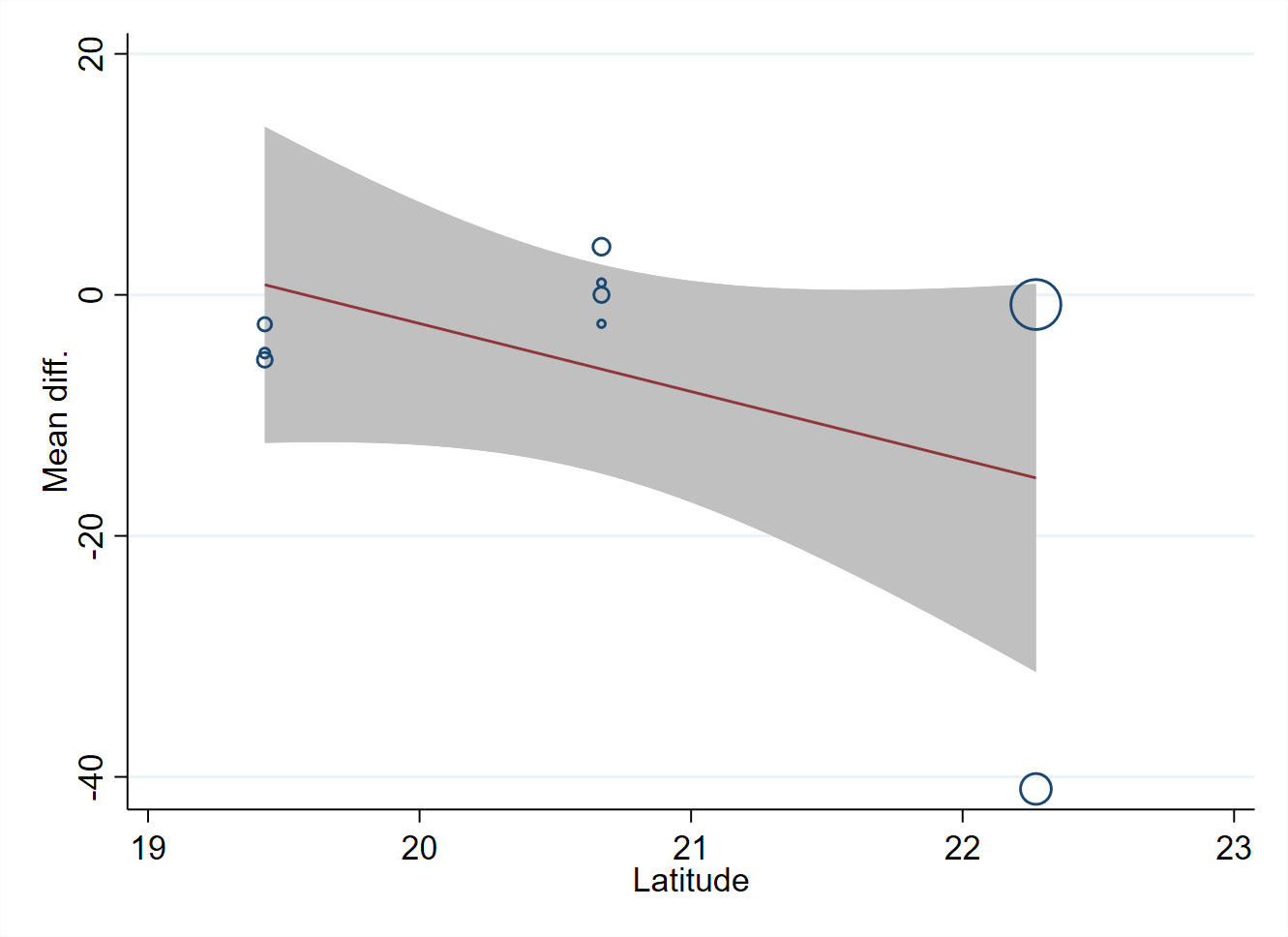

Supplemental Figure 2, panel B.
