## Supplemental Table 1 for "Looking for crumbs in the obesity forest: anti-obesity interventions in the Mexican population. History, and systematic review with Meta-Analysis"

| **Drugs (No diabetic patients)**  **DBP (Reference: placebo)** | | | |
| --- | --- | --- | --- |
| **Form1** | 0.000 (-5.117, 5.117) | 1.673 (-4.596, 7.942) | -0.256 (-5.380, 4.869) |
| 0.000 (-5.117, 5.117) | **Form2** | 1.673 (-4.597, 7.943) | -0.256 (-5.381, 4.870) |
| -1.673 (-7.942, 4.596) | -1.673 (-7.943, 4.597) | **Liraglutide** | -1.929 (-5.541, 1.684) |
| 0.256 (-4.869, 5.380) | 0.256 (-4.870, 5.381) | 1.929 (-1.684, 5.541) | **Plc** |

Supplemental table 1. Network meta-analysis results matrix. Estimates of the effect of treatments (standardized mean differences with 95%CI) relative to placebo (Plc). The Form1 and Form2 are described in the text, they are not approved by FDA.
