## Supplemental Table 2 for "Looking for crumbs in the obesity forest: anti-obesity interventions in the Mexican population. History, and systematic review with Meta-Analysis"

| **Drugs (Diabetic patients)**  **Glucose (Reference: metformin)** | | | |
| --- | --- | --- | --- |
| **Glim** | 0.000 (-0.466, 0.466) | 1.332 ( 0.587, 2.077) | -0.174 (-0.644, 0.296) |
| 0.000 (-0.466, 0.466) | **Glim+Met** | 1.332 ( 0.581, 2.083) | -0.174 (-0.654, 0.306) |
| -1.332 (-2.077, -0.587) | -1.332 (-2.083, -0.581) | **Insulin** | -1.506 (-2.084, -0.928) |
| 0.174 (-0.296, 0.644) | 0.174 (-0.306, 0.654) | 1.506 ( 0.928, 2.084) | **Met** |
| **DBP (Reference: metformin)** | | | |
| **Diac+Met** | -0.289 (-1.522, 0.944) | -0.796 (-2.036, 0.444) | -0.289 (-1.430, 0.851) |
| 0.289 (-0.944, 1.522) | **Glim** | -0.507 (-0.980, -0.033) | 0.000 (-0.469, 0.469) |
| 0.796 (-0.444, 2.036) | 0.507 ( 0.033, 0.980) | **Glim+Met** | 0.507 ( 0.020, 0.994) |
| 0.289 (-0.851, 1.430) | 0.000 (-0.469, 0.469) | -0.507 (-0.994, -0.020) | **Met** |

Supplemental table 2. Network meta-analysis results matrix. Estimates of the effect of treatments (standardized mean differences with 95%CI) relative to metformin in patients with diabetes. The Form1 and Form2 are described in the text, they are not approved by FDA.
